# Evidence for causal links between known modifiable risk factors and dementia: A systematic review of Mendelian randomization studies

**DOI:** 10.1101/2024.08.23.24312475

**Authors:** Roopal Desai, Amber John, Emma Anderson, Jean Stafford, Natalie L. Marchant, Georgina Charlesworth, Verena Zuber, Joshua Stott

## Abstract

**Background:** We aimed to systematically review the evidence for causal associations between the known modifiable risk factors and dementia based on Mendelian randomization (MR) studies.

**Method:** Five databases were searched from inception to April 2024 investigating the association between the twelve risk factors identified in the Lancet Commission and dementia. Evaluable analyses were categorised into one of four levels (robust, probable, suggestive, insufficient) based on estimate significance level and concordance of direction of effect between main and sensitivity analyses.

**Results:** 47 articles were included representing 160 separate analyses (136 unique; 104 evaluable) for ten risk factors and six dementia outcomes. There were no valid analyses for air pollution and traumatic brain injury. Of the unique and evaluable analyses over half (59.3%) were evaluated as providing ‘insufficient’ evidence of causal links. There was no evidence that genetically predicted liability to hearing loss was associated with dementia and limited genetic evidence for social contact. Evidence for education, obesity, depression, alcohol consumption and physical activity was inconclusive. There was probable evidence that smoking was protective against dementia risk however this may be an artefact of survivor bias. The two risk factors with the strongest genetic evidence for links with dementia were diabetes (probable evidence) and blood pressure (probable and suggestive evidence).

**Conclusion:** Genetic evidence for eight of the risk factors examined was insufficient or inconclusive. However, the null findings should be interpreted in the light of the biases inherent to MR studies. The strongest genetic evidence supported a causal link between diabetes and dementia.

## 1. Introduction

Dementia is a neurodegenerative set of diseases which typically affect people aged 65 and over. Although dementia is not an inevitable consequence of aging, age is the strongest predictor for dementia and as lifespans increase across the globe so will the number of people living with dementia. In 2019 the number of people globally living with dementia was estimated to be in excess of 55 million^1^ and this figure is forecast to rise to an estimated figure of 79 million by 2030 and further increase to 139 million by 2050^2^. The cost of dementia is great and is borne formally through the state and informally by individuals affected and their carers. Taking both these costs together the global cost of dementia was estimated as 1.3 trillion USD in 2019^1^. To date the progress on developing effective pharmaceutical interventions to halt or dimmish the disease has been slow and as a result research has increasingly expanded to identifying modifiable life-style risk factors for dementia^3^.

The Lancet Commission report on dementia prevention, intervention and care^3,4^ extensively reviewed the evidence on modifiable risk factors for dementia. The authors identified nine key risk factors, divided into three life stage categories, early (age<45 years), mid (age 45-65) and late (age>65). Less education was found to be an early life risk factor for dementia, with hearing loss, hypertension and obesity representing mid-life risks, and late-life factors including smoking, depression, physical inactivity, social isolation and diabetes. These nine risk factors were further extended in 2020^3^ to twelve by inclusion of the mid-life risk factor of excessive alcohol consumption and traumatic brain injury and the late-life risk factor of air pollution.

For each risk factor the Lancet Commission calculated the population attributable fraction (PAF); a statistic used to estimate the proportion of disease cases in a population that can be attributed to the specific risk factor. Combining the PAFs for all 12 risk factors, the overall PAF was estimated to be 40%. In other words, findings from the Lancet Commission have been interpreted as meaning that elimination of the 12 of the risk factors would result in a reduction of 40% of incident dementia. However, this interpretation is based on a key assumption in the PAF calculation of a *causal relationship* between the risk factor and the outcome. Given that the majority of the evidence in the Lancet Commission report came from observational studies, it is not possible to infer causation.

One approach to building evidence for causality in epidemiology is through triangulating results using different approaches^5^. Triangulation is the idea that different methodological approaches are vulnerable to distinct sources of biases. If two or more approaches aimed at answering the same question yield similar associations, then the evidence of a causal link is strengthened. Mendelian randomization (MR) is a method that has emerged in recent years as a powerful tool for identifying causal relationships between modifiable risk factors and diseases^6^. MR uses genetic variants as instrumental variables to test the causal effect of an exposure on an outcome. This approach takes advantage of the fact that genetic variants are randomly allocated at conception and are therefore less susceptible to the confounding and reverse causation biases common in observational studies. MR is subject to a different set of biases^5^ than traditional observational studies and thus an appropriate tool for examining evidence of causal links via triangulation.

In recent years, several MR studies have been conducted to investigate the relationship between modifiable risk factors and dementia. These studies have provided valuable insights into the potential modifiable risk factors for dementia and the potential impact of interventions targeting these risk factors. This systematic review aims to synthesise the findings of these MR studies and provide an overview of the evidence for links between known modifiable risk factors and dementia using MR.

## 2. Methods

### 2.1 Systematic search and study selection

The protocol for the current review was completed in advance and registered on PROSPERO (CRD42021254793). Five databases were searched from inception to April 2024. These databases were: Medline, Embase, PsycINFO, PubMed and Web of Science. Search terms relevant to MR (Mendelian randomization OR instrumental variable OR genetic instrument OR causal inference) were combined with search terms relevant to cognition or dementia (Alzheimer* OR dement* OR cognit* OR neurocognit* OR memory OR vascular dementia OR mild cognitive impairment OR MCI OR cognitive dysfunction OR cognition change OR frontotemporal dementia OR Lewy body dementia). After de-duplication, the remaining articles were subject to a title and abstract screen to identify relevant articles for full text inspection. Articles were assessed for inclusion based on predetermined inclusion and exclusion criteria. Articles were included if they were: MR studies assessing the association between a modifiable risk factor and dementia or dementia related outcome (e.g. age of onset, proxy risk); the modifiable risk factor was one of the twelve risk factors for dementia as identified by the Lancet Commission on Dementia^3^; available in English; and peer reviewed. Articles were excluded if they were: non-genetic studies or genetic studies other than MR; studies with outcomes that were not directly dementia-related (e.g. biomarker studies); review articles; and animal studies. Relevant reviews and all studies identified for inclusion were further subject to forward (citation searching) and backward searching (hand searching reference lists). One reviewer (RD) completed the primary search, title and abstract screen and full text screen and second reviewer (AJ) carried out an independent title and abstract and full text screen on 10% of the hit results at each stage. Inter-rater reliability was calculated for each stage of the screening process. Disagreements were resolved through discussion in consensus meetings and inter-rater reliability was calculated for each stage of the screening process.

### 2.2 Data extraction

Two reviewers (RD and AJ) independently extracted all the data (Supplementary Table 1), including; information on exposure and outcome, the genetic instrument, number of single nucleotide polymorphisms (SNP) *F*-statistic, the genome wide association study (GWAS) datasets used in the analysis, MR design (i.e. one-sample or two sample), the main MR estimate as reported by the study author and additional estimates obtained as part of sensitivity analyses.

### 2.3 Evaluation of evidence

Each MR estimate was evaluated for robustness of evidence for causality based on an established framework.^7^ One reviewer (RD) completed the evaluation on all estimates and another reviewer (EA) independently evaluated 20% of the estimates with disagreements resolved through discussion. As the evaluation was based on assessment of both the main analysis and at least one additional sensitivity analysis (e.g. MR Egger) if a study did not report any MR estimates in addition to their main finding, the estimate was categorised as ‘non-evaluable’. Where an estimate was deemed to be evaluable it was further categorised into one of four levels: ‘robust’, ‘probable’, ‘suggestive’, and ‘insufficient’.

*Robust genetic evidence* for causality was defined as the main MR estimate and all additional sensitivity estimates reported as significant, with concordant directions of effect. The threshold for significance was taken as the one the study authors used i.e., if a multiple testing corrected significance level was used that was taken as the significance threshold. If a corrected significance level was not reported then the standard threshold (*p*<0.05) was used.

*Probable genetic evidence* for causality was defined as at least one method, either the main or a sensitivity estimate, being significant (as defined by the study authors) with concordant directions of effect.

*Suggestive genetic evidence* for causality was defined as at least one method, either the main or a sensitivity estimate being significant, with non-concordant directions effect. In addition, in the situation where no sensitivity analysis was reported but the main MR result was reported as significant this was evaluated as ‘suggestive’ rather than ‘non-evaluable’. This was based on the conservative premise that had sensitivity analyses been conducted and the reported estimates were non-significant and non-concordant then the estimate would have been evaluated as ‘suggestive’.

*Insufficient genetic evidence* for causality was defined as all estimates being non-significant or the instrumental variable being deemed as not valid.

The number of estimates falling into each category of robustness evaluation was summed to create an aggregated value. To aggregate these values, only unique combinations of exposure and outcome datasets were included. That is, as the majority of GWAS datasets used in MR studies are publicly available, multiple study authors had conducted similar analyses using the same exposure and outcome GWAS datasets. Where multiple studies used the same exposure and outcome dataset combination, only one estimate was included in the aggregated results. In this situation, the included estimate was the one deemed to be evaluable. Where multiple analyses were evaluable then the study estimate included was the one with the highest quality rating.

### 2.4 Quality assessment

A quality assessment on all studies using a quality scoring system as proposed by Treur et al., (2021)^8^. This scoring system rates the quality of important aspects of MR studies including; phenotype measurement (sample size and quality of the exposure and outcome measurements); instrument strength (*p*-value threshold, number of SNPs, biological knowledge, *F*-statistic and % variance that the instrument explains). Each aspect given a quality rating which can be interpreted as a ‘low’, ‘moderate’ and ‘high’. An overall rating was achieved by consideration of key indicators. Two study authors (RD & J Stafford) independently rated each study with disagreements resolved in consensus meetings.

## 3. Results

### 3.1 Study selection

Initial database searches identified 3555 articles. An additional five studies were identified via citation searching. After removal of duplicates, 1711 articles were retained for title and abstract screening. At this stage 1605 were removed leaving 106 articles for full text inspection. Screening of the full texts in relation to inclusion and exclusion criteria resulted in 47 studies to be included. The PRISMA flow diagram of study selection is presented in Figure 1 and details of included studies are presented in Table 1. Inter-rater reliability for all stages was excellent (Cohen’s kappa >0.9).

**Figure 1:**
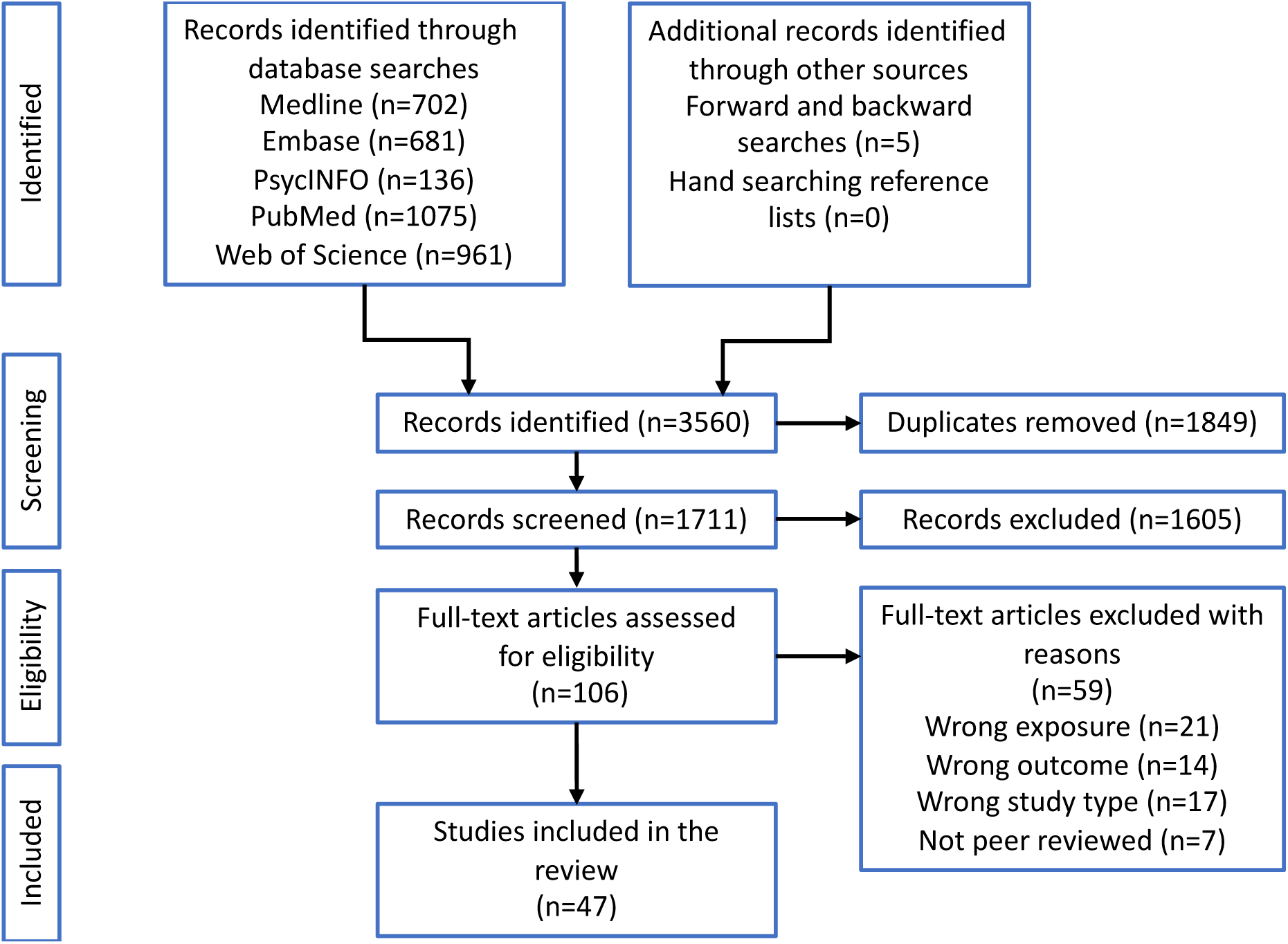
Flow diagram of study selection.

**Table 1.** Included studies and exposures and outcomes investigated.

| Study | Exposures investigated | Outcome | Method | Quality Assessment |
| --- | --- | --- | --- | --- |
| Abidin et al., (2021)<sup>1</sup> | Age related hearing loss | AD | Two-sample MR | Moderate |
| Anderson et al.,(2020)<sup>2</sup> | Educational attainment | AD | Two-sample MR | High |
| Andrews et al., (2020)<sup>3</sup> | Alcohol related exposures: alcohol consumption; alcohol dependence; AUDIT | AD; AAOS | Two-sample MR | High |
| Andrews et al., (2021)<sup>4</sup> | Alcohol related exposures: AUDIT, alcohol consumption<br>BMI<br>Depression<br>Educational attainment<br>Hearing difficulties<br>Hypertension related exposures: diastolic blood pressure; systolic blood pressure<br>Moderate to vigorous physical activity<br>Smoking related exposures: cigarettes per day; smoking initiation<br>Social isolation<br>Type 2 diabetes | AD | Two-sample MR | Moderate |
| Baumeister et al., (2020)<sup>5</sup> | Physical activity related exposures: average physical activity; vigorous physical activity | AD | Two-sample MR | High |
| Chen et al., (2022)<sup>6</sup> | Education related exposures: college or university degree; O-Levels/GCSEs or equivalents; no qualifications; age completed full time education<br>Obesity related exposures: BMI; body fat percentage; whole body fat-free mass<br>Physical activity related exposure: time spent using a computer | AD | Two-sample MR | High |
| Chen et al., (2023)<sup>7</sup> | Obesity related exposures: BMI; waist-hip ratio; waist-hip ratio adjusted for BMI | AD & AD by proxy | Two-sample MR | High |
| Desai et al., (2023)<sup>8</sup> | Alcohol related exposures: alcohol consumption<br>BMI<br>Depression<br>Educational attainment<br>Hearing difficulties<br>Hypertension related exposures: systolic blood pressure<br>Moderate to vigorous physical activity | AD & AD by proxy, AD; DLB; FTD | Two-sample MR | High |
|  | Smoking related exposures: lifetime smoking duration, heaviness, and cessation<br>Social isolation<br>Type 2 diabetes |  |  |  |
| Garfield et al., (2021)<sup>9</sup> | Diabetes related exposures: HbA1c; type 2 diabetes | AD | Two-sample MR | High |
| Harerimana et al., (2022)<sup>10</sup> | Depression | AD | Two-sample MR | Low |
| He et al., (2022)<sup>11</sup> | Physical activity related exposures: TV watching, computer use, driving | AD | Two-sample MR | Moderate |
| Hu et al., (2024)<sup>12</sup> | Depression | AD; VaD;<br>PDD; DLB;<br>FTD | Two-sample MR | High |
| Huang et al., (2023)<sup>13</sup> | Alcohol consumption<br>Education level<br>BMI<br>Diabetes related exposures: Type 2 diabetes; fasting glucose; fasting insulin; 2 h glucose; HbA1c<br>Hypertension related exposures: hypertension; systolic blood pressure; diastolic blood pressure<br>Smoking related exposures: smoking initiation; cigarettes per day | AD | Two-sample MR | Moderate |
| Korologou-Linden et al., (2022)<sup>14</sup> | Education related exposures: A-Levels; college degree<br>Obesity related exposure: whole body fat-free mass<br>Physical activity related exposure: self-reported moderate physical activity | AD | Two-sample MR | Moderate |
| Larsson et al., (2017)<sup>15</sup> | Alcohol consumption<br>Educational attainment<br>Diabetes related exposures: type 2 diabetes; fasting glucose; fasting insulin<br>Hypertension related exposures: diastolic blood pressure; systolic blood pressure<br>Obesity related exposures: BMI; waist to hip ratio adjusted BMI<br>Smoking related exposures: smoking quantity; smoking initiation; smoking cessation | AD | Two-sample MR | Moderate |
| Li et al., (2021)<sup>16</sup> | Obesity related exposures: BMI; waist to hip ratio; waist circumference; body fat percentage | AD | Two-sample MR | Moderate |
| Liao et al., (2022)<sup>17</sup> | Physical activity related exposures: fraction of acceleration >425 milli-gravities; overall acceleration average; moderate to vigorous physical activity; vigorous physical activity | AD | Two-sample MR | Low |
| Litkowski et al., (2023)<sup>18</sup> | Diabetes related exposures: type 2 diabetes; HbA1c; fasting glucose; fasting insulin | AD; all-cause dementia; VaD | One-sample MR | Moderate |
| Liu et al., (2022)<sup>19</sup> | Educational attainment | AD; AD by proxy; AD & AD by proxy | Two-sample MR | Low |
| Luo et al., (2023)<sup>20</sup> | Alcohol consumption<br>Blood<br>Education related exposures: number of years<br>Hypertension related exposures: diastolic blood pressure; systolic blood pressure<br>Smoking status<br>Type 2 diabetes | AD | Two-sample MR | High |
| Malik et al., (2021)<sup>21</sup> | BMI<br>HbA1c<br>Overall physical activity<br>Systolic blood pressure<br>Smoking index | Incident dementia | Two-sample MR | Moderate |
| Meng et al., (2022)<sup>22</sup> | Diabetes related exposures: type 2 diabetes; fasting glucose; insulin levels | AD | Two-sample MR | Low |
| Mitchell et al., (2020)<sup>23</sup> | Hearing impairment | AD | Two-sample MR | Low |
| Mukerjee et al., (2015)<sup>24</sup> | BMI | AD; dementia | One-sample MR | Moderate |
| Mulugeta et al., (2021)<sup>25</sup> | Adiposity related exposures: unfavourable metabolic profile for BMI | AD | One-sample MR<br>Two-sample MR | Moderate<br>Low |
| Ning et al., (2023)<sup>26</sup> | Air pollution related exposures: particulate matter 2.5 (PM<sub>2.5</sub>); particulate matter 10 (PM<sub>10</sub>); nitrogen dioxide (NO<sub>2</sub>); nitrogen oxide (NO<sub>x</sub>) | AD | Two-sample MR | Low |
| Nordesthaard et al., (2022)<sup>27</sup> | Smoking cumulative | All-cause dementia; AD; non-AD dementia | One-sample MR<br>Two-sample MR | Moderate<br>Moderate |
| Østergaard et al., (2015)<sup>28</sup> | BMI<br>Diabetes related exposures: fasting glucose; insulin resistance; type 2 diabetes<br>Education related exposures: length of education, completing university<br>Systolic blood pressure<br>Smoking related exposures: smoking quantity; smoking initiation | Alzheimer's Disease | Two-sample MR | Low |
| Ou et al., (2021)<sup>29</sup> | Hypertension related exposures: diastolic blood pressure systolic blood pressure; pulse pressure | AD | Two-sample MR | High |
| Pan et al., (2020)<sup>30</sup> | Diabetes related exposures: HbA1c; type 2 diabetes; fasting glucose; fasting insulin; homeostasis model assessment -B-cell function; homeostasis model assessment -insulin resistance | AD | Two-sample MR | Low |
| Raghavan et al., (2019)<sup>31</sup> | Educational attainment | AD | Two-sample MR | Low |
| Shen et al., (2021)<sup>32</sup> | Social isolation related exposures: regular pub attendance; regular gym attendance; regular religious group attendance; loneliness | AD; AD by proxy | Two-sample MR | Moderate |
| Sproviero et al., (2021)<sup>33</sup> | Hypertension related exposures: diastolic blood pressure; systolic blood pressure | AD | Two-sample MR | Moderate |
| Thomassen et al., (2020)<sup>34</sup> | Type 2 diabetes | AD | Two-sample MR | Moderate |
| Thorp et al., (2022)<sup>35</sup> | Alcohol related exposures: drinks per week<br>BMI<br>Depression<br>Education related exposures: educational attainment; cognitive component of educational attainment; non-cognitive component of educational attainment<br>Hearing impairment<br>Hypertension related exposures: diastolic blood pressure; systolic blood pressure<br>Loneliness<br>Physical activity<br>Smoking related exposures: smoking initiation; cigarettes per day<br>Type 2 diabetes | AD | Two-sample MR | High |
| Walter et al., (2016)<sup>36</sup> | Diabetes related exposures: type 2 diabetes; type 2 diabetes-adiposity; type 2 diabetes -beta cell function; type 2 diabetes – insulin sensitivity; type 2 diabetes – other biological factors | AD | Two-sample MR | Moderate |
| Wang et al., (2020)<sup>37</sup> | Educational attainment | AD | Two-sample MR | High |
| Wang et al., (2024)<sup>38</sup> | BMI | AD | Two-sample MR | High |
| Wu et al., (2021)<sup>39</sup> | Physical activity | AD | Two-sample MR | Moderate |
| Xue et al., (2023)<sup>40</sup> | Type 2 diabetes | AD | Two-sample MR | Moderate |
| Yang et al., (2021)<sup>41</sup> | Physical activity related exposures: sedentary behaviour (TV watching);<br>sedentary behaviour (computer use); sedentary behaviour (driving) | AD | Two-sample MR | High |
| Zhang et al., (2020)<sup>42</sup> | BMI<br>Educational attainment<br>Current smoking<br>Diabetes related exposures: type 2 diabetes; fasting insulin; fasting glucose | AD | Two-sample MR | Moderate |
| Zhang et al., (2022)<sup>43</sup> | Physical activity related exposures: overall activity; sedentary behaviour;<br>walking; moderate intensity activity | AD | Two-sample MR | Moderate |
| Zhou et al., (2019)<sup>44</sup> | Obesity related exposures: BMI; waist to hip ratio; waist circumference; waist<br>to hip ratio adjusted BMI | AD | Two-sample MR | High |
| Zhou et al., (2022)<sup>45</sup> | Diabetes related exposures: fasting insulin; insulin resistance | AD | Two-sample MR | Moderate |
| Zhu et al., (2023)<sup>46</sup> | Smoking cigarettes per day | AD | Two-sample MR | High |
| Zhuang et al., (2021)<sup>47</sup> | Obesity | AD | Two-sample MR | Low |

### 3.2 Description of studies

The 47 studies meeting inclusion criteria represented 240 unique MR associations for eleven of the Lancet Commission modifiable dementia risk factors exposure categories and nine different dementia outcomes. There were no MR studies examining traumatic brain injury and dementia risk.

#### Exposures

##### Early-life risk factor

Twenty five (10.4%) analyses for *education*-related exposures; educational attainment (n=15, 6.3%); college or university degree (n=3, 1.3%); O-Levels or equivalents (n=1, 0.4%); A-Levels or equivalent (n=1, 0.4%); no qualifications (n=1, 0.4%); age on completing education (n=1, 0.4%); length of education (n=1, 0.4%); the cognitive component of education (n=1, 0.4%); the non-cognitive component of education (n=1, 0.4%).

##### Mid-life risk factors

Thirty-four (14.2%) *adiposity*-related exposures including: body mass index (BMI) (n=18, 7.5%); body fat percentage (n=2, 0.8%); whole body fat-free mass (n=2, 0.8%); waist to hip ratio adjusted BMI (n=5, 2.1%); waist to hip ratio (n=2, 0.8%); waist circumference (n=2, 0.8%); unfavourable metabolic profile for BMI (n=2, 0.8%) and obesity (n=1, 0.4%). Fifteen (6.3%) analyses for *alcohol*-related exposures including: alcohol consumption (n=10, 4.2%); alcohol dependence (n=2, 0.8%); alcohol use disorders identification test (AUDIT) (n=2, 0.8%) and drinks per week (n=1, 0.4%).

Thirty (12.5%) *blood pressure*-related exposures including: diastolic blood pressure (n=10, 4.2%); systolic blood pressure (n=13, 5.4%), pulse pressure (n=4, 1.7%) and hypertension (n=3, 1.6%).

Seven *hearing loss*-related exposures including: age-related hearing loss (n=4, 1.7%); hearing difficulties (n=1, 0.4%) and hearing impairment (n=2, 0.8%).

##### Late-life risk factors

Four (1.7%) *air pollution*-related exposures including: particulate matter 2.5 (n=1, 0.4%); particulate matter 10 (n=1, 0.4%); nitrogen dioxide (n=1, 0.4%) and nitrogen oxides (n=1, 0.4%).

Ten (4.2%) *depression* exposures including: self-report depression and hospital admission (n=10, 4.2%).

Fifty-three (22.1%) *diabetes*-related exposures including: type 2 diabetes (n=25, 10.4%); type 2 diabetes adiposity (n=1, 0.4%); type 2 diabetes insulin sensitivity (n=1, 0.4%); type 2 diabetes other biological factors (n=1, 0.4%); type 2 diabetes β cell function (n=1, 0.4%); HbAC1 (n=6, 2.5%); 2 hour postprandial glucose (n=3, 1.3%); fasting glucose (n=5, 2.1%); fasting insulin (n=5, 2.1%); homeostasis β cell function (n=1, 0.4%); homeostasis insulin resistance (n=1, 0.4%); insulin resistance (n=2, 0.8%), insulin levels (n=1, 0.4%).

Twenty-two *physical activity*-related exposures (9.2%) including: overall physical activity (n=7, 2.9%); vigorous physical activity (n=5, 2.1%); moderate to vigorous physical activity (n=1, 0.4%); moderate physical activity (n=1, 0.4%); walking (n=1, 0.4%); sedentary behaviour (n=1, 0.4%); sedentary behaviour -computer use (n=2, 0.8%); sedentary behaviour -TV watching (n=1, 0.4%); sedentary behaviour -driving (n=1, 0.4%); fraction of accelerations >425 milli-gravities (n=1, 0.4%); overall acceleration (n=1, 0.4%).

Twenty-five (10.4%) *smoking*-related exposures including: current smoking (n=1, 0.4%); lifetime smoking (n=2, 0.8%); smoking initiation (n=6, 2.5%); smoking quantity (n=2, 0.8%); smoking cessation (n=1, 0.4%); smoking cumulative (n=4, 1.7%); smoking index (n=1, 0.4%); smoking cigarettes per day (n=6, 2.5%); smoking regularly (n=2, 0.8%).

Fifteen (6.3%) *social contact*-related exposures including: social isolation (n=1, 0.4%); regular social club or pub attendance (n=2, 0.8%); regular sport club or gym attendance (n=2, 0.8%); regular religious group attendance (n=2, 0.8%); loneliness (n=8, 3.3%).

##### Outcome categories

The dementia outcomes included: Alzheimer’s disease (AD) (n=149, 62.1%); AD by proxy (n=14, 5.8%), maternal AD (n=14, 5.8%), paternal AD (n=14, 5.8%), combined AD and AD by proxy (n=8, 3.3%), AD age of onset (n=3, 1.3%), all cause dementia (n=11, 4.6%), non-AD dementia (n=1, 0.4%), dementia with Lewy bodies (DLB) (n=10, 4.2%), frontotemporal dementia (FTD) (n=11, 4.6%), Parkinson’s disease dementia (PDD) (n=1, 0.4%) and vascular dementia (VaD) (n=4, 1.7%).

### 3.3 Quality Assessment

18 (38.3%) studies were rated ‘high’ quality, 19 (40.4%) studies were rated of ‘moderate’ quality and 11 studies (23.4%) were rated ‘low’ quality (see Supplementary Materials for details). The 11 studies given a ‘low’ quality rating were removed before the evidence was evaluated.

### 3.4 Evaluation of evidence

#### Early-life risk factor

Education: there were 17 unique analyses investigating the relationship between education in early life and dementia outcomes (Table 2). Of these, one (5.9%) was graded as ‘robust’ and nine (52.9%) as ‘probable’ evidence of a causal relationship. All of the studies graded as ‘robust’ or ‘probable’ indicated that genetically determined greater educational attainment was protective against developing AD aside from one^9^ which used a proxy measure for AD and found the opposite effect. Two (8.7%) studies were graded as ‘suggestive’ and five (21.7%) were graded as providing insufficient evidence.

**Table 2.**
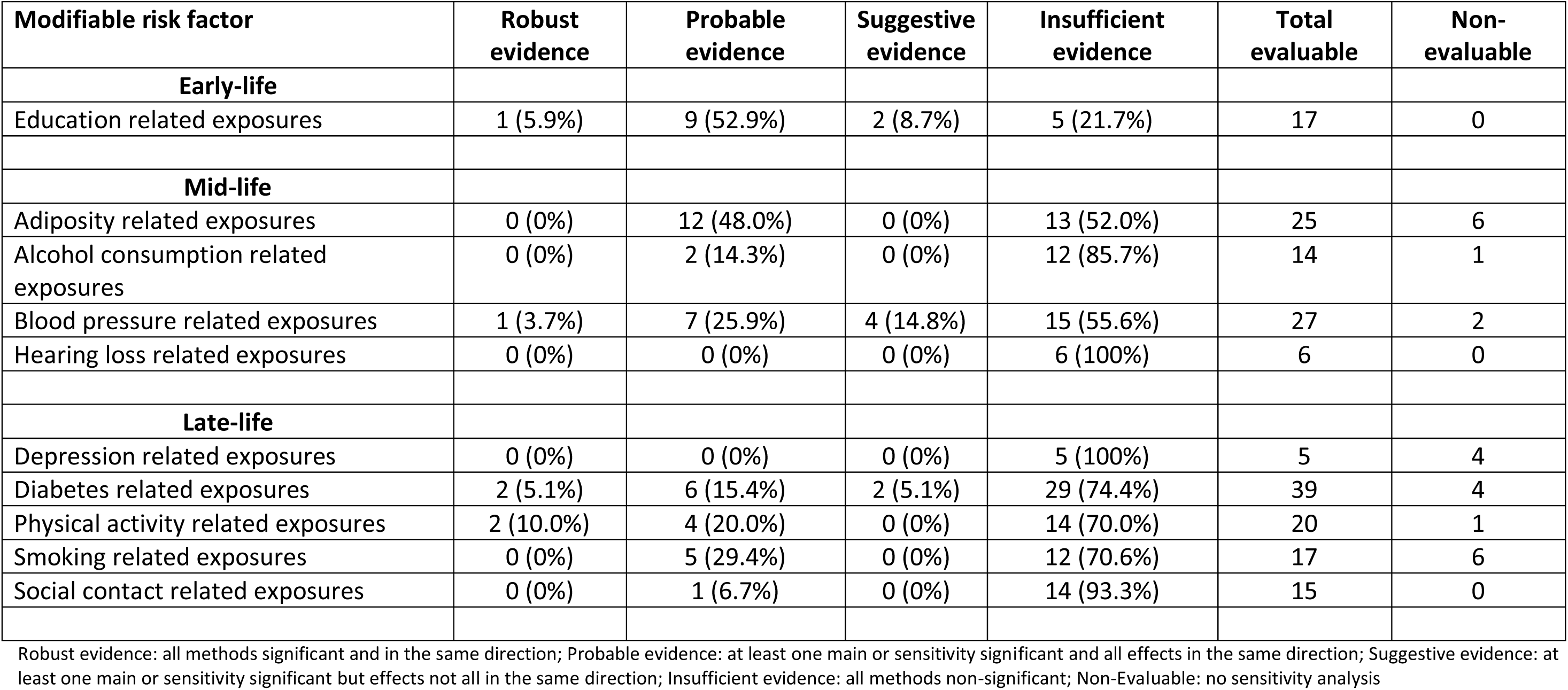
Number and percentage of ‘high’ or ‘moderate’ quality, unique Mendelian randomization analyses per category by modifiable risk factor.

#### Mid-life risk factors

Adiposity related risk factors: After exclusion of six non-evaluable analyses there were 25 unique analyses for the mid-life risk factor of adiposity and dementia outcomes. Of these none were graded as ‘robust’ and 12 (48.0%) were graded as ‘probable’. Three of these analyses indicated that genetically determined higher adiposity-related phenotype was associated with greater risk of dementia and nine analyses indicated a protective relationship. No analyses were graded as ‘suggestive’ and 13 were deemed as providing insufficient evidence.

Alcohol: After exclusion of one non-evaluable analyses there were 14 unique MR analysis for the mid-life risk factor of alcohol consumption and dementia outcomes. None of these were graded as ‘robust’ and two (14.3%) were graded as ‘probable’. The direction of these effects was not aligned. One analysis indicted that genetically determined greater alcohol consumption was associated with an earlier age of AD onset^10^ and another indicated that genetically determined higher alcohol dependence was associated with a later age of AD onset^10^. The remaining analyses (85.7%) were graded as ‘insufficient’.

Blood pressure related exposures: After exclusion of two non-evaluable analyses there were 27 unique MR analysis for the mid-life risk factor of high blood pressure. One (3.7%) was graded as ‘robust’ indicating that higher diastolic blood pressure was protective against AD^9^. Seven (25.0%) were graded as ‘probable’ with non-concordant directions of effects. Four (14.8%) analyses indicated ‘suggestive’ evidence of casual relationships between genetically determined higher blood pressure and AD. Fifteen analyses were graded as ‘insufficient’.

Hearing loss: There were six unique analyses for the association between age related hearing loss and dementia outcomes. All six (100%) of these were deemed as providing ‘insufficient’ evidence for causal links.

#### Late-life risk factors

Depression: There were five unique analyses for the late-life risk factor of depression. All five were graded as ‘insufficient’.

Diabetes: After exclusion of four non-evaluable analyses there were 39 unique analyses for diabetes-related exposures and dementia outcomes. Of these two (5.1%) were graded as providing ‘robust’ evidence of causal relationships between type 2 diabetes and an increased risk of all cause dementia and VaD. Six (15.4%) were graded as ‘probable’ with all analyses indicating diabetes related metabolic increased the risk of dementia. Two (5.1%) analyses were graded as ‘suggestive’ and the remining 29 (74.4%) were graded as ‘insufficient’.

Physical activity: After exclusion of one non-evaluable analysis there were 20 unique analyses for physical activity related exposures. Of these two (10.0%) were graded as ‘robust’, four (20.0%) graded as ‘probable’ and 14 (70.0%) ‘insufficient’. Of the two ‘robust’ analyses, genetically determined higher levels of physical activity in the form of walking was found to be protective against AD^11^. However, when sedentary behaviour as measured by computer use was used as the exposure the analysis indicated that genetically determined higher levels of sedentary behaviour were protective against AD^12^. Of the four analyses graded as ‘probable’ the direction of effects was contradictory depending on the exposure. Two analyses found that genetically determined higher levels of overall physical activity or lower levels of sedentary behaviour^13^ were associated with a decreased risk of AD and the other two found that greater levels of self-reported moderate activity^14^ or lower levels of sedentary behaviour^15^ increased the risk of dementia.

Smoking: After exclusion of six non-evaluable analyses there were seventeen unique analyses examining the relationship between smoking related exposures and dementia outcomes. Of these none were graded as ‘robust’, five (42.9%) were graded as ‘probable’ and 12 (57.1%) were graded as ‘insufficient’. All five analyses graded as ‘probable’ found that genetically determined higher levels of smoking was protective against AD.

Social contact: There were 15 unique analyses examining the relationship between social contact and dementia. Of these none were graded as ‘robust’ one (6.7%) was deemed as providing ‘probable’ evidence of a causal relationship. This analysis indicated that genetically determined higher levels of gym attendance was protective against AD as measured by proxy^16^. Fourteen (93.3%) were graded as providing ‘insufficient’ evidence of causal links.

For all the exposures aside from education related and depression related exposures the bulk (>50%) of evidence was graded as providing ‘insufficient’ evidence of causal links.

## 4. Discussion

We aimed to summarise the MR evidence for causal relationships between twelve modifiable risk factors and dementia outcomes. Studies were identified for eleven risk factors; there were no studies relevant traumatic brain injury. Of the unique, evaluable analyses identified and graded in this study over half (67.6%) indicated *insufficient* evidence for causal links between the exposures and dementia outcomes. For hearing loss and depression, the limited available evidence was categorised entirely as insufficient and indeed the only factors for which evidence was not predominantly insufficient were education. However, although both probable and robust genetic evidence of relationships were available for education, high blood pressure, and physical activity, findings for the *direction* of effect (risk factor or protective factor) were contradictory, possibly explained by methodological limitations in MR studies such as use of proxy outcomes^17^ or poor concept validity for exposures. The only risk factor for which robust and probable relationships were consistently in the direction of risk was diabetes. Contradictory evidence of probable relationships was also identified for alcohol consumption and obesity. Contrary to expectations, smoking was found to be a protective factor in all five analyses categorised as probable, potentially explained by survivor bias^18^. The single analyses showing probable relationships between social contact and dementia outcomes indicated a protective nature of social contact.

The protective effect of education is generally considered a robust and consistent epidemiological finding based on the triangulation of evidence from multiple study designs with different sources of bias and it is unexpected to find an MR analysis in the opposite direction. However, education as a risk (rather than a protective) factor was only evident when a *proxy* measure of AD was used as the outcome. This proxy measure is reliant on self-report of parental AD status and therefore may not be an objective way to measure AD risk^19^. It is also of note that when educational attainment was divided into the distinct concepts of the cognitive (i.e. intelligence) and non-cognitive (e.g. conscientiousness) components it was only the cognitive component that was found to be protective against dementia risk^20^. It is unclear if intelligence is as modifiable as education and as such whether increasing the number of years in education will be protective in and of itself.

The results for the risk factors of alcohol consumption, obesity, physical activity and high blood pressure were inconclusive with the direction of effect varying across different analyses. This was the case even when proxy measures of AD were not considered as outcomes.

There was some evidence that smoking may have a mitigating effect on dementia risk. However, this result needs to be interpreted with caution because there is strong evidence to link smoking to overall negative health outcomes and premature all-cause mortality^21^. In addition, the results that are observed in MR studies may be an artefact of survivor bias^22^. Individuals who smoke may die earlier of other smoking-related diseases and therefore may not survive long enough to develop an age-related disease. As such what appears to be a link with dementia may be an association with longevity.

One risk factor namely type 2 diabetes and related biomarkers had the most evidence of causal links to dementia outcomes with the direction of effect aligned over analyses. This was the risk factor that had comparatively the strongest overall evidence to be causally linked to dementia.

### Limitations and future research

MR studies for dementia face several challenges, notably in identifying valid instrumental variables, construct validity, survivor bias and lack of subtype specific analyses. For IVs to be valid there must be a biologically plausible pathway between the IV exposure and outcome. The most notable case of this limitation is that of the one study investigating the effect of air pollution on dementia^23^. These analyses were all deemed non-evaluable due to concerns over the lack of biological plausibility of the IV^24^.Studies may be limited on how well the exposure or outcome in question is operationalised. Exposures such as physical activity and social contact were susceptible to construct validity bias. Physical inactivity in one study^15^ was assessed as time spent on a computer when this may also be a measure of mental activity or work. Likewise in another study social contact was operationalised as regular gym attendance^16^ when this could also be a measure of physical activity rather than social contact. Diagnostic challenges further hinder MR studies; diagnostic overlap between dementia subtypes and minimal phenotyping (e.g., reliance on proxy phenotypes) can lead to misclassification and bias, affecting the accuracy and reliability of study findings. Survivor bias also complicates MR studies, as dementia typically manifests later in life, meaning individuals exposed to certain risk factors may be at risk from dying earlier in life from a non-dementia related illness. Additionally, the availability of detailed data on dementia subtypes, particularly VaD, DLB and FTD is limited, complicating subtype-specific analyses. Another limitation of MR studies is that there is no agreed standardised way to conduct MR analyses. Heterogeneity may be introduced at various stages of the analysis pipeline including at the instrument selection, clumping and pruning stages^25^. One important limitation of this review is that here we only consider the risk factors for dementia as identified by the Lancet Commission. MR studies in the field of dementia research have focused on a broad range of modifiable risk factors from coffee consumption^26^ to cholesterol levels^27^ as well as many biomarkers^28^. Future reviews should focus on evaluating the evidence from all these non-Lancet Commission risk modifiable factors in order to provide a comprehensive review of all the MR evidence in the field of dementia.

#### Conclusion

Currently, the evidence-base for causal relationships between ten of the Lancet commission risk factors for dementia is insufficient when the evidence from MR studies is considered. However, null results may be due to the limitations of MR studies. More studies using a range of different study designs are needed in order to triangulate results and allow us to better understand the nature of the relationships between lifestyle factors and dementia. Further reviews of the MR literature need to be completed to synthesis evidence from emerging risk factors (i.e. non Lancet Commission risk factors) coming from MR studies and dementia related outcomes. In addition, there is a need to adapt MR methods to account for systematic forms of bias.

## Data Availability

All data produced in the present work are contained in the manuscript

