## SupplementaryFile for "Evidence for causal links between known modifiable risk factors and dementia: A systematic review of Mendelian randomization studies"

Supplementary Table1: Results of included studies investigating modifiable risk factors and dementia

| Modifiable Risk Factor | Study | Exposure (GWAS dataset/consortium) | Outcome (GWAS dataset/consortium) | N SNPs | *F*-statistic | IVW Estimate |
|---|---|---|---|---|---|---|
| **Early Life** | | | | | | |
| **Education related exposures** | Anderson et al.,(2020)[2] | Educational attainment (Okbay et al., 2016[48]) | Alzheimer's Disease (Lambert et al., 2013[49]) | 142 | 43.5 | OR=0.62 95% CI:0.51;0.77*** |
| | Andrews et al., (2021)[4] | Educational attainment (Lee et al., 2018[50]) | Alzheimer's Disease (Kunkle et al., 2019[51]) | 478 | 55.9 | β=-0.44 SE=0.07*** |
| | Chen et al., (2022)[6] | College or University degree (MRC-IEU consortium) | Alzheimer's Disease (Kunkle et al., 2019[51]) | 211 | 46.7 | OR=0.47 95% CI:0.34;0.67*** |
| | | O-Levels or equivalent (MRC-IEU consortium) | Alzheimer's Disease (Kunkle et al., 2019[51]) | 18 | 35.6 | OR=0.28 95% CI:0.09;0.89* |
| | | No qualifications (MRC-IEU consortium) | Alzheimer's Disease (Kunkle et al., 2019[51]) | 78 | 42.1 | OR=4.01 95% CI:1.95;8.21** |
| | | Age completed education (MRC-IEU consortium) | Alzheimer's Disease (Kunkle et al., 2019[51]) | 37 | 36 | OR=0.76 95% CI:0.48;1.20* |
| | Desai et al., (2023)[8] | Educational attainment (Okbay et al., 2016[48]) | Alzheimer's Disease and Alzheimer's Disease by Proxy (Bellenguez et al., 2023[52]) | 70 | 45.8 | OR=1.93 95% CI:1.05;3.54 |
| | | Educational attainment (Okbay et al., 2016[48]) | Dementia with Lewy Bodies (Chia et al., 2021[53]) | 67 | 45.5 | OR=1.99 95% CI:0.24;16.23 |
| | | Educational attainment (Okbay et al., 2016[48]) | Frontotemporal Dementia (Ferrari et al., 2014[54]) | 71 | 46 | OR=2.84 95% CI:0.24;33.57 |
| | Huang et al., (2023)[13] | Educational attainment (Lee et al., 2018[50]) | Alzheimer's Disease (Kunkle et al., 2019[51]) | 262 | NR | OR=0.70 95% CI:0.60;0.81** |
| | | Educational attainment (Lee et al., 2018[50]) | Alzheimer's Disease by Proxy - Maternal history (Marioni et al., 2018[55]) | 262 | NR | OR=1.31 95% CI:1.10;1.57** |
| | | Educational attainment (Lee et al., 2018[50]) | Alzheimer's Disease by Proxy - Paternal history (Marioni et al., 2018[55]) | 262 | NR | OR=1.21 95% CI:0.94;1.56 |
| | Korologou-Linden et al., (2022)[14] | A-Levels (UK Biobank) | Alzheimer's Disease (Lambert et al., 2013[49]) | 77 | >30 | OR=0.82 95% CI:0.73;0.91*** |
| | | College degree (UK Biobank) | Alzheimer's Disease (Lambert et al., 2013[49]) | 241 | >30 | OR=0.82 95% CI:0.76;0.88*** |

| Modifiable Risk Factor | Study | Exposure (GWAS dataset/consortium) | Outcome (GWAS dataset/consortium) | N SNPs | *F*-statistic | IVW Estimate |
|---|---|---|---|---|---|---|
| | Larsson et al., (2017)[15] | Educational attainment (Okbay et al., 2016[48]) | Alzheimer's Disease (Lambert et al., 2013[49]) | 152 | 5.7 | OR=0.89 95% CI:0.84;0.93*** |
| | Liu et al., (2022)[19] | Educational attainment (Okbay et al., 2016[48]) | Alzheimer's Disease (Kunkle et al., 2019[51]) | 159 | NR | OR=0.68 95% CI:0.57;0.81** |
| | | Educational attainment (Okbay et al., 2016[48]) | Alzheimer's Disease by proxy (Marioni et al., 2018[55]) | 147 | NR | OR=1.09 95% CI:1.00;1.19 |
| | | Educational attainment (Okbay et al., 2016[48]) | Alzheimer's Disease by proxy (Schwartzentruber et al., 2021[56]) | 159 | NR | OR=1.88 95% CI:1.62;2.18*** |
| | | Educational attainment (Okbay et al., 2016[48]) | Alzheimer's Disease & Alzheimer's Disease by proxy (Marioni et al., 2018[55]) | 147 | NR | OR=0.99 95% CI:0.91;1.07 |
| | | Educational attainment (Okbay et al., 2016[48]) | Alzheimer's Disease & Alzheimer's Disease by proxy (Jansen et al., 2019[57]) | 155 | NR | OR=0.96 95% CI:0.93;0.98** |
| | | Educational attainment (Okbay et al., 2016[48]) | Alzheimer's Disease & Alzheimer's Disease by proxy (Schwartzentruber et al., 2021[56]) | 159 | NR | OR=1.21 95% CI:1.08;1.36** |
| | Luo et al., (2023)[20] | Educational attainment (Okbay et al., 2016[48]) | Alzheimer's Disease (Bellenguez et al., 2022[52]) | 3201 | >10 | OR=0.83 95% CI:0.79;0.87** |
| | | Educational attainment (Okbay et al., 2016[48]) | Alzheimer's Disease by proxy (Bellenguez et al., 2022[52]) | 3197 | >10 | OR=1.06 95% CI:1.01;1.10 |
| | Østergaard et al., (2015)[28] | Length of education (Rietveld et al., 2013[58]) | Alzheimer's Disease (Lambert et al., 2013[49]) | 1 | NR | OR=0.71 95% CI:0.48;1.06 |
| | | Completing university (Rietveld et al., 2013[58]) | Alzheimer's Disease (Lambert et al., 2013[49]) | 2 | NR | OR=0.95 95% CI:0.67;1.34 |
| | Raghavan et al., (2019)[31] | Educational attainment (Okbay et al., 2016[48]) | Alzheimer's Disease (Lambert et al., 2013[49]) | 1271 | NR | OR=0.63 95% CI:0.54;0.74*** |
| | Thorp et al., (2022)[35] | Educational attainment (Lee et al., 2018[50]) | Alzheimer's Disease (Lambert et al., 2013[49]) | 271 | 48.8 | OR=0.66 95% CI:0.55;0.78*** |
| | | Cognitive component of educational attainment (Lee et al., 2018[50] and Savage et al., 2018[59]) | Alzheimer's Disease (Lambert et al., 2013[49]) | 121 | 42.4 | OR=0.85 95% CI:0.78;0.92** |

| Modifiable Risk Factor | Study | Exposure (GWAS dataset/consortium) | Outcome (GWAS dataset/consortium) | N SNPs | *F*-statistic | IVW Estimate |
|---|---|---|---|---|---|---|
| | | Non cognitive component of educational attainment (Lee et al., 2018[50] and Savage et al., 2018[59]) | Alzheimer's Disease (Lambert et al., 2013[49]) | 28 | 36.9 | OR=0.94 95% CI:0.82;1.08 |
| | Wang et al., (2020)[37] | Educational attainment (Okbay et al., 2016[48]) | Alzheimer's Disease (Lambert et al., 2013[49]) | 64 | NR | OR=0.59 95% CI:0.45;0.77** |
| | Zhang et al., (2020)[42] | Educational attainment (Lee et al., 2018[50]) | Alzheimer's Disease (Lambert et al., 2013[49]) | 248 | NR | OR=0.67 95% CI:0.57;0.80*** |
| **Mid-Life** | | | | | | |
| **Alcohol consumption related exposures** | Andrews et al., (2020)[3] | Alcohol consumption (Liu et al., 2019[60]) | Alzheimer's Disease (Lambert et al., 2013[49]) | 44 | 70.8 | OR=0.96 95% CI:0.74;1.25 |
| | | Alcohol dependence (Walter et al., 2018[61]) | Alzheimer's Disease (Lambert et al., 2013[49]) | 20 | 24.6 | OR=0.93 95% CI:0.93;1.04 |
| | | AUDIT (Sanchez-Roige et al., 2019[62]) | Alzheimer's Disease (Lambert et al., 2013[49]) | 11 | 59.3 | OR=0.45 95% CI:0.12;1.75 |
| | | Alcohol consumption (Liu et al., 2019[60]) | Alzheimer's Disease Age of Onset Survival (Huang et al., 2017[63]) | 44 | 70.8 | HR=2.02 95% CI:1.42;2.87*** |
| | | Alcohol dependence (Walter et al., 2018[61]) | Alzheimer's Disease Age of Onset Survival (Huang et al., 2017[63]) | 20 | 24.6 | HR=0.93 95% CI:0.87;0.99* |
| | | AUDIT (Sanchez-Roige et al., 2019[62]) | Alzheimer's Disease Age of Onset Survival (Huang et al., 2017[63]) | 11 | 59.3 | $\beta$=0.26 SE=0.18 |
| | Andrews et al., (2021)[4] | AUDIT (Sanchez-Roige et al., 2019[62]) | Alzheimer's Disease (Kunkle et al., 2019[51]) | 12 | 58.2 | $\beta$=0.59 SE=0.29* |
| | | Alcohol consumption (Liu et al., 2019[60]) | Alzheimer's Disease (Kunkle et al., 2019[51]) | 86 | 69.4 | $\beta$=0.28 SE=0.26 |
| | Desai et al., (2023)[8] | Alcohol consumption (Evangelou et al., 2019)[64] | Alzheimer's Disease and Alzheimer's Disease by Proxy (Bellenguez et al., 2023[52]) | 72 | 45.8 | OR=1.19 95% CI:0.85;1.66 |
| | | Alcohol consumption (Evangelou et al., 2019)[64] | Dementia with Lewy Bodies (Chia et al., 2021[53]) | 69 | 58 | OR=0.60 95% CI:0.18;2.03 |

| Modifiable Risk Factor | Study | Exposure (GWAS dataset/consortium) | Outcome (GWAS dataset/consortium) | N SNPs | *F*-statistic | IVW Estimate |
|---|---|---|---|---|---|---|
| | | Alcohol consumption (Evangelou et al., 2019)[64] | Frontotemporal Dementia (Ferrari et al., 2014[54]) | 65 | 61.9 | OR=1.54 95% CI:1.20;1.97* |
| | Huang et al., (2023)[13] | Alcohol consumption (Liu et al., 2019[60]) | Alzheimer's Disease (Kunkle et al., 2019[51]) | 32 | NR | OR=1.22 95% CI:0.83;1.78 |
| | | Alcohol consumption (Liu et al., 2019[60]) | Alzheimer's Disease by Proxy - Maternal history (Marioni et al., 2018[55]) | 32 | NR | OR=1.27 95% CI:0.84;1.94 |
| | | Alcohol consumption (Liu et al., 2019[60]) | Alzheimer's Disease by Proxy - Paternal history (Marioni et al., 2018[55]) | 32 | NR | OR=0.95 95% CI:0.53;1.69 |
| | Larsson et al., (2017)[15] | Alcohol consumption (Jorgenson et al., 2017[65]) | Alzheimer's Disease (Lambert et al., 2013[49]) | 3 | NR | OR=0.72 95% CI:0.5;1.04 |
| | Luo et al., (2023)[20] | Alcohol consumption (Liu et al., 2019[60]) | Alzheimer's Disease (Bellenguez et al., 2022[52]) | 77 | >10 | OR=0.87 95% CI:0.70;1.07 |
| | | Alcohol consumption (Liu et al., 2019[60]) | Alzheimer's Disease by proxy (Bellenguez et al., 2022[52]) | 78 | >10 | OR=0.93 95% CI:0.74;1.15 |
| | Thorp et al., (2022)[35] | Drinks per week (Liu et al., 2019[60]) | Alzheimer's Disease (Lambert et al., 2013[49]) | 29 | 76.4 | OR=1.08 95% CI:0.71;1.65 |
| **Blood pressure related exposures** | Andrews et al., (2021)[4] | Diastolic blood pressure (Evangelou et al., 2018[66]) | Alzheimer's Disease (Kunkle et al., 2019[51]) | 450 | 82.6 | $\beta$=-0.001 SE=0.004* |
| | | Systolic blood pressure (Evangelou et al., 2018[66]) | Alzheimer's Disease (Kunkle et al., 2019[51]) | 435 | 78.1 | $\beta$=-0.005 SE=0.003 |
| | Desai et al., (2023)[8] | Systolic blood pressure (UKBiobank) | Alzheimer's Disease and Alzheimer's Disease by Proxy (Bellenguez et al., 2023[52]) | 219 | 45.4 | OR=0.90 95% CI:0.82;0.99* |
| | | Systolic blood pressure (UKBiobank) | Dementia with Lewy Bodies (Chia et al., 2021[53]) | 213 | 45.6 | OR=1.12 95% CI:0.82;1.52 |
| | | Systolic blood pressure (UKBiobank) | Frontotemporal Dementia (Ferrari et al., 2014[54]) | 212 | 45.1 | OR=0.95 95% CI:0.67;1.37 |
| | Huang et al., (2023)[13] | Hypertension (UK Biobank) | Alzheimer's Disease (Kunkle et al., 2019[51]) | 143 | NR | OR=1.45 95% CI:1.03;2.03* |

| Modifiable Risk Factor | Study | Exposure (GWAS dataset/consortium) | Outcome (GWAS dataset/consortium) | N SNPs | *F*-statistic | IVW Estimate |
|---|---|---|---|---|---|---|
| | | Hypertension (UK Biobank) | Alzheimer's Disease by Proxy - Maternal history (Marioni et al., 2018[55]) | 138 | NR | OR=1.50 95% CI:0.98;2.30 |
| | | Hypertension (UK Biobank) | Alzheimer's Disease by Proxy - Paternal history (Marioni et al., 2018[55]) | 138 | NR | OR=2.24 95% CI:1.26;4.00* |
| | | Diastolic blood pressure (UK Biobank) | Alzheimer's Disease (Kunkle et al., 2019[51]) | 400 | NR | OR=0.95 95% CI:0.67;1.37 |
| | | Diastolic blood pressure (UK Biobank) | Alzheimer's Disease by Proxy - Maternal history (Marioni et al., 2018[55]) | 380 | NR | OR=1.00 95% CI:0.99;1.01 |
| | | Diastolic blood pressure (UK Biobank) | Alzheimer's Disease by Proxy - Paternal history (Marioni et al., 2018[55]) | 380 | NR | OR=0.99 95% CI:0.99;1.00 |
| | | Systolic blood pressure (UK Biobank) | Alzheimer's Disease (Kunkle et al., 2019[51]) | 398 | NR | OR=0.99 95% CI:0.98;1.00 |
| | | Systolic blood pressure (UK Biobank) | Alzheimer's Disease by Proxy - Maternal history (Marioni et al., 2018[55]) | 374 | NR | OR=0.99 95% CI:0.98;1.00 |
| | | Systolic blood pressure (UK Biobank) | Alzheimer's Disease by Proxy - Paternal history (Marioni et al., 2018[55]) | 374 | NR | OR=0.98 95% CI:0.96;0.99* |
| | | Pulse Pressure (UK Biobank) | Alzheimer's Disease (Kunkle et al., 2019[51]) | 343 | NR | OR=1.00 95% CI:0.98;1.01 |
| | | Pulse Pressure (UK Biobank) | Alzheimer's Disease by Proxy - Maternal history (Marioni et al., 2018[55]) | 326 | NR | OR=0.99 95% CI:0.98;1.01 |
| | | Pulse Pressure (UK Biobank) | Alzheimer's Disease by Proxy - Paternal history (Marioni et al., 2018[55]) | 326 | NR | OR=0.99 95% CI:0.98;1.01 |
| | Larsson et al., (2017)[15] | Diastolic blood pressure (Hoffmann et al., 2017[67]) | Alzheimer's Disease (Lambert et al., 2013[49]) | 105 | 11 | OR=0.96 95% CI:0.79;1.16 |

| Modifiable Risk Factor | Study | Exposure (GWAS dataset/consortium) | Outcome (GWAS dataset/consortium) | N SNPs | *F*-statistic | IVW Estimate |
|---|---|---|---|---|---|---|
| | | Systolic blood pressure (Hoffmann et al., 2017[67]) | Alzheimer's Disease (Lambert et al., 2013[49]) | 93 | 15 | OR=0.94 95% CI:0.77;1.14 |
| | Luo et al., (2023)[20] | Diastolic blood pressure (Evangelou et al., 2018[66]) | Alzheimer's Disease (Bellenguez et al., 2022[52]) | 216 | >10 | OR=1.02 95% CI:0.96;1.09 |
| | | Diastolic blood pressure (Evangelou et al., 2018[66]) | Alzheimer's Disease by proxy (Bellenguez et al., 2022[52]) | 215 | >10 | OR=0.98 95% CI:0.93;1.03 |
| | | Systolic blood pressure (Evangelou et al., 2018[66]) | Alzheimer's Disease (Bellenguez et al., 2022[52]) | 268 | >10 | OR=0.89 95% CI:0.81;0.98* |
| | | Systolic blood pressure (Evangelou et al., 2018[66]) | Alzheimer's Disease by proxy (Bellenguez et al., 2022[52]) | 268 | >10 | OR=0.85 95% CI:0.78;0.92* |
| | Malik et al., (2021)[21] | Systolic blood pressure (UKBiobank) | Incident Dementia (UK Biobank) | 460 | NR | OR=1.31 95% CI:1.05;1.60 |
| | Østergaard et al., (2015)[28] | Systolic blood pressure (Ehret et al., 2011[68]) | Alzheimer's Disease (Lambert et al., 2013[49]) | 24 | NR | OR=0.75 95% CI:0.62;0.91 |
| | Ou et al., (2021)[29] | Diastolic blood pressure (Evangelou et al., 2018[66]) | Alzheimer's Disease (Kunkle et al., 2019[51]) | 398 | NR | OR=0.99 95% CI:0.98;1.00 |
| | | Systolic blood pressure (Evangelou et al., 2018[66]) | Alzheimer's Disease (Kunkle et al., 2019[51]) | 400 | NR | OR=1.00 95% CI:0.99;1.01 |
| | | Pulse pressure (Evangelou et al., 2018[66]) | Alzheimer's Disease (Kunkle et al., 2019[51]) | 343 | NR | OR=1.00 95% CI:0.98;1.01 |
| | Sproviero et al., (2021)[33] | Diastolic blood pressure (Evangelou et al., 2018[66]) | Alzheimer's Disease (Lambert et al., 2013[49]) | 56 | NR | OR=0.95 95% CI:0.80;1.10 |
| | | Systolic blood pressure (Evangelou et al., 2018[66]) | Alzheimer's Disease (Lambert et al., 2013[49]) | 63 | NR | OR=0.91 95% CI:0.83;0.99 |
| | Thorp et al., (2022)[35] | Diastolic blood pressure (Evangelou et al., 2018[66]) | Alzheimer's Disease (Lambert et al., 2013[49]) | 363 | 80.6 | OR=0.99 95% CI:0.98;1.00 |
| | | Systolic blood pressure (Evangelou et al., 2018[66]) | Alzheimer's Disease (Lambert et al., 2013[49]) | 343 | 77.9 | OR=1.00 95% CI:0.99;1.00 |
| **Hearing loss related exposures** | Abidin et al., (2021)[1] | Age related hearing loss (Wells et al., 2019[69]) | Alzheimer's Disease (Kunkle et al., 2019[51]) | 34 | NR | $\beta$=-0.26 SE=0.30 |
| | Andrews et al., (2021)[4] | Hearing difficulties (Wells et al., 2019[69]) | Alzheimer's Disease (Kunkle et al., 2019[51]) | 40 | 43.1 | $\beta$=-0.13 SE=0.27 |

| Modifiable Risk Factor | Study | Exposure (GWAS dataset/consortium) | Outcome (GWAS dataset/consortium) | N SNPs | *F*-statistic | IVW Estimate |
|---|---|---|---|---|---|---|
| | Desai et al., (2023)[8] | Age related hearing loss (Kalra et al., 2020)[70] | Alzheimer's Disease and Alzheimer's Disease by Proxy (Bellenguez et al., 2023[52]) | 30 | 43.4 | OR=1.07 95% CI:0.85;1.33 |
| | | Age related hearing loss (Kalra et al., 2020)[70] | Dementia with Lewy Bodies (Chia et al., 2021[53]) | 29 | 43.4 | OR=0.75 95% CI:0.30;1.88 |
| | | Age related hearing loss (Kalra et al., 2020)[70] | Frontotemporal Dementia (Ferrari et al., 2014[54]) | 26 | 26 | OR=0.89 95% CI:0.30;2.66 |
| | Mitchell et al., (2020)[23] | Hearing impairment (Wells et al., 2019[69]) | Alzheimer's Disease (Lambert et al., 2013[49]) | 35 | NR | $\beta$=0.04 SE=0.06 |
| | Thorp et al., (2022)[35] | Hearing impairment (Wells et al., 2019[69]) | Alzheimer's Disease (Lambert et al., 2013[49]) | 40 | 40.9 | OR=0.89 95% CI:0.48;1.74 |
| **Obesity related exposures** | Andrews et al., (2021)[4] | Body mass index (Yengo et al., 2018[71]) | Alzheimer's Disease (Kunkle et al., 2019[51]) | 505 | 76.6 | $\beta$=-0.03 SE=0.045 |
| | Chen et al., (2022)[6] | Body mass index (MRC-IEU consortium) | Alzheimer's Disease (Kunkle et al., 2019[51]) | 371 | 65.5 | OR=0.79 95% CI:0.58;1.10 |
| | | Body fat percentage (MRC-IEU consortium) | Alzheimer's Disease (Kunkle et al., 2019[51]) | 220 | 59.9 | OR=0.62 95% CI:0.38;1.00 |
| | | Whole body fat-free mass (MRC-IEU consortium) | Alzheimer's Disease (Kunkle et al., 2019[51]) | 471 | 89.1 | OR=0.79 95% CI:0.69;0.90** |
| | Chen et al., (2023)[7] | Body mass index (Pulit et al., 2019[72]) | Alzheimer's Disease (Jansen et al., 2019[57]) | 472 | >10 | OR=1.04 95% CI:1.02;1.05*** |
| | | Waist to hip ratio (Pulit et al., 2019[72]) | Alzheimer's Disease (Jansen et al., 2019[57]) | 296 | >10 | OR=1.03 95% CI:1.01;1.05** |
| | | Waist to hip ratio adjusted body mass index (Pulit et al., 2019[72]) | Alzheimer's Disease (Jansen et al., 2019[57]) | 293 | >10 | OR=0.98 95% CI:0.98;1.01 |
| | Desai et al., (2023)[8] | Waist to hip ratio adjusted body mass index (Pulit et al., 2019[72]) | Alzheimer's Disease and Alzheimer's Disease by Proxy (Bellenguez et al., 2023[52]) | 542 | 55.3 | OR=0.87 95% CI:0.82;0.96* |
| | | Waist to hip ratio adjusted body mass index (Pulit et al., 2019[72]) | Dementia with Lewy Bodies (Chia et al., 2021[53]) | 538 | 55.5 | OR=0.75 95% CI:0.57;0.99* |
| | | Waist to hip ratio adjusted body mass index (Pulit et al., 2019[72]) | Frontotemporal Dementia (Ferrari et al., 2014[54]) | 526 | 55.3 | OR=0.75 95% CI:0.58;0.97* |

| Modifiable Risk Factor | Study | Exposure (GWAS dataset/consortium) | Outcome (GWAS dataset/consortium) | N SNPs | *F*-statistic | IVW Estimate |
|---|---|---|---|---|---|---|
| | Huang et al., (2023)[13] | Body mass index (Yengo et al., 2018[71]) | Alzheimer's Disease (Kunkle et al., 2019[51]) | 715 | NR | OR=0.88 95% CI:0.83;0.94* |
| | | Body mass index (Yengo et al., 2018[71]) | Alzheimer's Disease by Proxy - Maternal history (Marioni et al., 2018[55]) | 715 | NR | OR=0.92 95% CI:0.86;0.99* |
| | | Body mass index (Yengo et al., 2018[71]) | Alzheimer's Disease by Proxy - Paternal history (Marioni et al., 2018[55]) | 715 | NR | OR=0.93 95% CI:0.84;1.02 |
| | Korologou-Linden et al., (2022)[14] | Whole body fat-free mass (UK Biobank) | Alzheimer's Disease (Lambert et al., 2013[49]) | 521 | >30 | OR=0.79 95% CI:0.70;0.89*** |
| | Larsson et al., (2017)[15] | Body mass index (Locke et al., 2015[73]) | Alzheimer's Disease (Lambert et al., 2013[49]) | 76 | 17 | OR=1.05 95%C CI:0.91;1.21 |
| | | Waist to hip ratio adjusted body mass index (Shungin et al., 2015[74]) | Alzheimer's Disease (Lambert et al., 2013[49]) | 38 | 17 | OR=1.18 95% CI:0.97;1.45 |
| | Li et al., (2021)[16] | Body mass index (Pulit et al., 2019[72]) | Alzheimer's Disease (Jansen et al., 2019[57]) | 305 | NR | OR=1.04 95% CI:1.01;1.05* |
| | | Waist to hip ratio (Pulit et al., 2019[72]) | Alzheimer's Disease (Jansen et al., 2019[57]) | 201 | NR | OR=1.02 95% CI:1.00;1.04 |
| | | Waist circumference (Shungin et al., 2015[74]) | Alzheimer's Disease (Jansen et al., 2019[57]) | 38 | NR | OR=1.03 95% CI:1.00;1.07* |
| | | Body fat percentage (Lu et al., 2016[75]) | Alzheimer's Disease (Jansen et al., 2019[57]) | 9 | NR | OR=1.00 95% CI:0.95;1.06 |
| | Luo et al., (2023)[20] | Body mass index (Yengo et al., 2018[71]) | Alzheimer's Disease (Bellenguez et al., 2022[52]) | 440 | >10 | OR=0.94 95% CI:0.87;1.01 |
| | | Body mass index (Yengo et al., 2018[71]) | Alzheimer's Disease by proxy (Bellenguez et al., 2022[52]) | 440 | >10 | OR=0.89 95% CI:0.83;0.95 |
| | Malik et al., (2021)[21] | Body mass index (UKBiobank) | Incident dementia (UKBiobank) | 941 | NR | OR=0.88 95% CI:0.72;1.08 |
| | Mukerjee et al., (2015)[24] | Body mass index (Alzheimer's Disease Genetics Consortium) | Alzheimer's Disease (Alzheimer's Disease Genetics Consortium) | 31 | NR | OR=0.95 95% CI:0.90;1.01 |
| | | Body mass index (Health and Retirement Study) | Dementia (Health and Retirement Study) | 29 | NR | OR=1.00 95% CI:0.75;1.32 |

| Modifiable Risk Factor | Study | Exposure (GWAS dataset/consortium) | Outcome (GWAS dataset/consortium) | N SNPs | *F*-statistic | IVW Estimate |
|---|---|---|---|---|---|---|
| | | Body mass index (Genetic and Environmental Risk for AD consortium) | Alzheimer's Disease (Genetic and Environmental Risk for AD consortium) | NR | NR | OR=0.96 95% CI:0.87;1.07 |
| | Mulugeta et al., (2021)[25] | Unfavourable metabolic profile for body mass index (UKBiobank) | Alzheimer's Disease (UKBiobank) | 82 | NR | OR=0.89 95% CI:0.60;1.32 |
| | | Unfavourable metabolic profile for body mass index (UKBiobank) | Alzheimer's Disease (UKBiobank) | 76 | NR | OR=0.89 95% CI:0.43;1.84 |
| | Østergaard et al., (2015)[28] | Body mass index (Speliotes et al., 2010[76]) | Alzheimer's Disease (Lambert et al., 2013[49]) | 49 | NR | OR=0.99 95% CI:0.80;1.19 |
| | Thorp et al., (2022)[35] | Body mass index (Yengo et al., 2018[71]) | Alzheimer's Disease (Lambert et al., 2013[49]) | 418 | 74.8 | OR=0.83 95% CI:0.64;1.07 |
| | Wang et al., (2024)[38] | Body mass index (Howe et al., 2022[77]) | Alzheimer's Disease (Kunkle et al., 2019[51]) | 42 | >10 | $\beta$=-0.037 95% CI:-0.078;0.004 |
| | Zhang et al., (2020)[42] | Body mass index (UKBiobank) | Alzheimer's Disease (Lambert et al., 2013[49]) | 360 | NR | OR=0.90 95% CI:0.82;1.00 |
| | Zhou et al., (2019)[44] | Body mass index (GIANT consortium) | Alzheimer's Disease (Lambert et al., 2013[49]) | 62 | 92.3 | OR=1.10 95% CI:0.90;1.34 |
| | | Waist to hip ratio (GIANT consortium) | Alzheimer's Disease (Lambert et al., 2013[49]) | 23 | 88.1 | OR=1.05 95% CI:0.76;1.45 |
| | | Waist circumference (GIANT consortium) | Alzheimer's Disease (Lambert et al., 2013[49]) | 12 | 65.5 | OR=0.97 95% CI:0.59;1.61 |
| | | Waist to hip ratio adjusted body mass index (GIANT consortium) | Alzheimer's Disease (Lambert et al., 2013[49]) | 36 | 108.2 | OR=1.12 95% CI:0.89;1.41 |
| | Zhuang et al., (2021)[47] | Obesity (GIANT consortium) | Alzheimer's Disease (Lambert et al., 2013[49]) | 14 | NR | OR=0.97 95% CI:0.89;1.06 |
| **Late-life** | | | | | | |
| **Air Pollution related exposures** | Ning et al., (2023)[26] | Particulate Matter 2.5 (UK Biobank) | Alzheimer's Disease (Kunkle et al., 2019[51]) | 7 | >10 | OR=1.77 95% CI:0.90;3.49 |
| | | Particulate Matter 10 (UK Biobank) | Alzheimer's Disease (Kunkle et al., 2019[51]) | 19 | >10 | OR=1.93 95% CI:1.03;3.59* |
| | | Nitrogen Dioxide (UK Biobank) | Alzheimer's Disease (Kunkle et al., 2019[51]) | 5 | >10 | OR=0.70 95% CI:0.24;1.98 |

| Modifiable Risk Factor | Study | Exposure (GWAS dataset/consortium) | Outcome (GWAS dataset/consortium) | N SNPs | *F*-statistic | IVW Estimate |
|---|---|---|---|---|---|---|
| | | Nitrogen Oxide (UK Biobank) | Alzheimer's Disease (Kunkle et al., 2019[51]) | 8 | >10 | OR=2.05 95% CI:0.86;4.87 |
| **Depression related exposures** | Andrews et al., (2021)[4] | Depression (Howard et al., 2019[78]) | Alzheimer's Disease (Kunkle et al., 2019[51]) | 84 | 43.5 | β=-0.15 SE=0.07* |
| | Desai et al., (2023)[8] | Depression (Howard et al., 2019[78]) | Alzheimer's Disease and Alzheimer's Disease by Proxy (Bellenguez et al., 2023[52]) | 76 | 42.8 | OR=1.02 95% CI:0.92;1.13 |
| | | Depression (Howard et al., 2019[78]) | Dementia with Lewy Bodies (Chia et al., 2021[53]) | 71 | 42.2 | OR=0.99 95% CI:0.64;1.55 |
| | | Depression (Howard et al., 2019[78]) | Frontotemporal Dementia (Ferrari et al., 2014[54]) | 78 | 41.9 | OR=1.00 95% CI:0.66;1.52 |
| | Harerimana et al., (2022)[10] | Depression (Howard et al., 2019[78]) | Alzheimer's Disease (Jansen et al., 2019[57]) | 115 | NR | β=0.029 SE=0.01* |
| | Hu et al., (2024)[12] | Depression (Howard et al., 2019[78]) | Alzheimer's Disease (GWAS ID: ieu-b-5067) | 48 | >10 | OR=1.00 95% CI:0.99;1.00 |
| | | Depression (Howard et al., 2019[78]) | Vascular Dementia (GWAS ID: finn-b-F5_VASCDEM) | 47 | >10 | OR=2.13 95% CI:1.25;3.64* |
| | | Depression (Howard et al., 2019[78]) | Parkinson's Disease Dementia (GWAS ID: finn-b-PD_DEMENTIA) | 47 | >10 | OR=0.59 95% CI:0.20;1.72 |
| | | Depression (Howard et al., 2019[78]) | Dementia with Lewy Bodies (Chia et al., 2021[53]) | 44 | >10 | OR=1.01 95% CI:0.60;1.70 |
| | | Depression (Howard et al., 2019[78]) | Frontotemporal Dementia (Van Deerlin et al., 2010[79]) | 23 | >10 | OR=1.48 95% CI:0.47;4.63 |
| | Thorp et al., (2022)[35] | Depression (Howard et al., 2019[78]) | Alzheimer's Disease (Lambert et al., 2013[49]) | 42 | 38.7 | OR=0.82 95% CI:0.67;1.01 |
| | Andrews et al., (2021)[4] | Type 2 diabetes (Xue et al., 2018[80]) | Alzheimer's Disease (Kunkle et al., 2019[51]) | 117 | 76.7 | β=-0.02 SE=0.02 |
| | Desai et al., (2023)[8] | Type 2 diabetes (Scott et al., 2017[81]) | Alzheimer's Disease and Alzheimer's Disease by Proxy (Bellenguez et al., 2023[52]) | 39 | 62.2 | OR=1.01 95% CI:0.96;1.05 |
| | | Type 2 diabetes (Scott et al., 2017[81]) | Dementia with Lewy Bodies (Chia et al., 2021[53]) | 38 | 62.5 | OR=1.02 95% CI:0.86;1.22 |

| Modifiable Risk Factor | Study | Exposure (GWAS dataset/consortium) | Outcome (GWAS dataset/consortium) | N SNPs | *F*-statistic | IVW Estimate |
|---|---|---|---|---|---|---|
| **Diabetes related exposures** | | Type 2 diabetes (Scott et al., 2017[81]) | Frontotemporal Dementia (Ferrari et al., 2014[54]) | 38 | 62.5 | OR=1.02 95% CI:0.86;1.22 |
| | Garfield et al., (2021)[9] | Haemoglobin A1c (Wheeler et al., 2017[82]) | Alzheimer's Disease (UK Biobank) | 51 | 164.6 | OR=1.09 95% CI:0.42;2.83 |
| | | Type 2 diabetes (Mahajan et al., 2018[83]) | Alzheimer's Disease (UK Biobank) | 157 | 27.4 | OR=1.15 95% CI:0.87;1.52 |
| | Huang et al., (2023)[13] | Type 2 diabetes (UK Biobank) | Alzheimer's Disease (Kunkle et al., 2019[51]) | 37 | NR | OR=0.74 95% CI:0.11;4.73 |
| | | Type 2 diabetes (UK Biobank) | Alzheimer's Disease by Proxy - Maternal history (Marioni et al., 2018[55]) | 34 | NR | OR=2.73 95% CI:0.59;12.62 |
| | | Type 2 diabetes (UK Biobank) | Alzheimer's Disease by Proxy - Paternal history (Marioni et al., 2018[55]) | 34 | NR | OR=0.29 95% CI:0.02;4.23 |
| | | Fasting Glucose (Chen et al., 2021[84]) | Alzheimer's Disease (Kunkle et al., 2019[51]) | 64 | NR | OR=1.08 95% CI:0.84;1.38 |
| | | Fasting Glucose (Chen et al., 2021[84]) | Alzheimer's Disease by Proxy - Maternal history (Marioni et al., 2018[55]) | 64 | NR | OR=0.86 95% CI:0.57;1.37 |
| | | Fasting Glucose (Chen et al., 2021[84]) | Alzheimer's Disease by Proxy - Paternal history (Marioni et al., 2018[55]) | 64 | NR | OR=0.89 95% CI:0.62;1.28 |
| | | Fasting Insulin (Chen et al., 2021[84]) | Alzheimer's Disease (Kunkle et al., 2019[51]) | 35 | NR | OR=1.06 95% CI:0.74;1.52 |
| | | Fasting Insulin (Chen et al., 2021[84]) | Alzheimer's Disease by Proxy - Maternal history (Marioni et al., 2018[55]) | 33 | NR | OR=0.98 95% CI:0.78;1.23 |
| | | Fasting Insulin (Chen et al., 2021[84]) | Alzheimer's Disease by Proxy - Paternal history (Marioni et al., 2018[55]) | 33 | NR | OR=0.82 95% CI:0.42;1.58 |
| | | 2 hr postprandial glucose (Chen et al., 2021[84]) | Alzheimer's Disease (Kunkle et al., 2019[51]) | 12 | NR | OR=1.04 95% CI:0.91;1.19 |

| Modifiable Risk Factor | Study | Exposure (GWAS dataset/consortium) | Outcome (GWAS dataset/consortium) | N SNPs | *F*-statistic | IVW Estimate |
|---|---|---|---|---|---|---|
| | | 2 hr postprandial glucose (Chen et al., 2021[84]) | Alzheimer's Disease by Proxy - Maternal history (Marioni et al., 2018[55]) | 10 | NR | OR=1.04 95% CI:0.87;1.24 |
| | | 2 hr postprandial glucose (Chen et al., 2021[84]) | Alzheimer's Disease by Proxy - Paternal history (Marioni et al., 2018[55]) | 10 | NR | OR=1.02 95% CI:0.80;1.30 |
| | | Haemoglobin A1c (Chen et al., 2021[84]) | Alzheimer's Disease (Kunkle et al., 2019[51]) | 68 | NR | OR=1.15 95% CI:0.83;1.59 |
| | | Haemoglobin A1c (Chen et al., 2021[84]) | Alzheimer's Disease by Proxy - Maternal history (Marioni et al., 2018[55]) | 66 | NR | OR=0.93 95% CI:0.68;1.25 |
| | | Haemoglobin A1c (Chen et al., 2021[84]) | Alzheimer's Disease by Proxy - Paternal history (Marioni et al., 2018[55]) | 66 | NR | OR=1.09 95% CI:0.70;1.69 |
| | Larsson et al., (2017)[15] | Type 2 diabetes (Morris et al., 2012[85]) | Alzheimer's Disease (Lambert et al., 2013[49]) | 50 | 62 | OR=1.02 95% CI:0.97;1.07 |
| | | Fasting glucose (Scott et al., 2017[81]) | Alzheimer's Disease (Lambert et al., 2013[49]) | 36 | 72 | OR=1.14 95% CI:0.99;1.32 |
| | | Fasting insulin (Scott et al., 2017[81]) | Alzheimer's Disease (Lambert et al., 2013[49]) | 19 | 34 | OR=1.13 95% CI:0.85;1.51 |
| | Litkowski et al., (2023)[18] | Type 2 diabetes (Mahajan et al., 2022[86]) | All Cause Dementia (European Million Veteran Cohort) | 330 | 740.4 | OR=1.07 95% CI:1.05;1.08*** |
| | | Type 2 diabetes (Mahajan et al., 2022[86]) | Vascular Dementia (European Million Veteran Cohort) | 330 | 740.4 | OR=1.11 95% CI:1.07;1.15*** |
| | | Type 2 diabetes (Mahajan et al., 2022[86]) | Alzheimer's Disease (European Million Veteran Cohort) | 330 | 740.4 | OR=1.06 95% CI:1.02;1.09*** |
| | | Type 2 diabetes (Mahajan et al., 2022[86]) | All Cause Dementia (African Million Veteran Cohort) | 330 | 57.3 | OR=1.06 95% CI:1.02;1.10* |
| | | Type 2 diabetes (Mahajan et al., 2022[86]) | Vascular Dementia (African Million Veteran Cohort) | 330 | 57.3 | OR=1.11 95% CI:1.04;1.19* |
| | | Type 2 diabetes (Mahajan et al., 2022[86]) | Alzheimer's Disease (African Million Veteran Cohort) | 330 | 57.3 | OR=1.12 95% CI:1.02;1.23* |

| Modifiable Risk Factor | Study | Exposure (GWAS dataset/consortium) | Outcome (GWAS dataset/consortium) | N SNPs | *F*-statistic | IVW Estimate |
|---|---|---|---|---|---|---|
| | | Type 2 diabetes (Mahajan et al., 2022[86]) | All Cause Dementia (Hispanic Million Veteran Cohort) | 330 | 61.8 | OR=1.04 95% CI:0.98;1.10 |
| | | Type 2 diabetes (Mahajan et al., 2022[86]) | Vascular Dementia (Hispanic Million Veteran Cohort) | 330 | 61.8 | OR=1.09 95% CI:0.96;1.23 |
| | | Type 2 diabetes (Mahajan et al., 2022[86]) | Alzheimer's Disease (Hispanic Million Veteran Cohort) | 330 | 61.8 | OR=0.94 95% CI:0.83;1.07 |
| | Luo et al., (2023)[20] | Type 2 diabetes (Vujkovic et al., 2020[87]) | Alzheimer's Disease (Bellenguez et al., 2022[52]) | 357 | >10 | OR=1.02 95% CI:0.98;1.05 |
| | | Type 2 diabetes (Vujkovic et al., 2020[87]) | Alzheimer's Disease by proxy (Bellenguez et al., 2022[52]) | 353 | >10 | OR=1.00 95% CI:0.97;1.03 |
| | Malik et al., (2021)[21] | Haemoglobin A1c (UK Biobank) | Incident Dementia (UK Biobank) | 176 | NR | OR=1.24 95% CI: 0.82;1.88 |
| | Meng et al., (2022)[22] | Type 2 diabetes (Mahajan et al., 2018[83]) | Alzheimer's Disease (Lambert et al., 2013[49]) | 37 | NR | OR=1.34 95% CI:1.05;1.70 |
| | | Fasting glucose (Wessel et al., 2015[88]) | Alzheimer's Disease (Lambert et al., 2013[49]) | 7 | NR | OR=1.57 95% CI:1.14;2.17 |
| | Østergaard et al., (2015)[28] | Fasting glucose (Scott et al., 2017[81]) | Alzheimer's Disease (Lambert et al., 2013[49]) | 36 | NR | OR=1.12 95% CI:0.97;1.30 |
| | | Insulin resistance (Scott et al., 2017[81]) | Alzheimer's Disease (Lambert et al., 2013[49]) | 10 | NR | OR=1.32 95% CI:0.88;1.98 |
| | | Type 2 diabetes (Morris et al., 2012[85]) | Alzheimer's Disease (Lambert et al., 2013[49]) | 49 | NR | OR=1.02 95% CI:0.97;1.07 |
| | Pan et al., (2020)[30] | Haemoglobin A1c (Wheeler et al., 2017[82]) | Alzheimer's Disease (Lambert et al., 2013[49]) | 36 | NR | OR=0.96 95% CI:0.69;1.32 |
| | | Type 2 diabetes (Morris et al., 2012[85]; Scott et al., 2017[81]) | Alzheimer's Disease (Lambert et al., 2013[49]) | 43 | NR | OR=1.02 95% CI:0.97;1.07 |
| | | Fasting glucose (Scott et al., 2017[81]) | Alzheimer's Disease (Lambert et al., 2013[49]) | 28 | NR | OR=1.33 95% CI:1.04;1.68* |
| | | Fasting insulin (Scott et al., 2017[81]) | Alzheimer's Disease (Lambert et al., 2013[49]) | 19 | NR | OR=1.24 95% CI:0.77;2.01 |
| | | Homeostasis model assessment -B-cell function (Dupuis et al., 2010[89]) | Alzheimer's Disease (Lambert et al., 2013[49]) | 6 | NR | OR=1.92 95% CI:1.15;3.21* |

| Modifiable Risk Factor | Study | Exposure (GWAS dataset/consortium) | Outcome (GWAS dataset/consortium) | N SNPs | *F*-statistic | IVW Estimate |
|---|---|---|---|---|---|---|
| | | Homeostasis model assessment -Insulin resistance (Dupuis et al., 2010[89]) | Alzheimer's Disease (Lambert et al., 2013[49]) | 2 | NR | OR=1.17 95% CI:0.40;3.37 |
| | Thomassen et al., (2020)[34] | Type 2 diabetes (Scott et al., 2017[81]) | Alzheimer's Disease (Lambert et al., 2013[49]) | 51 | NR | OR=1.04 95% CI:0.98;1.10 |
| | Thorp et al., (2022)[35] | Type 2 diabetes (Xue et al., 2018[80]) | Alzheimer's Disease (Lambert et al., 2013[49]) | 111 | 22.9 | OR=1.01 95% CI:0.96;1.06 |
| | Walter et al., (2016)[36] | Type 2 diabetes (Morris et al., 2012[85]) | Alzheimer's Disease (Lambert et al., 2013[49]) | 39 | NR | OR=1.01 95% CI:0.96;1.06 |
| | | Type 2 diabetes adiposity (Morris et al., 2012[85]) | Alzheimer's Disease (Lambert et al., 2013[49]) | 20 | NR | OR=0.93 95% CI:0.74;1.15 |
| | | Type 2 diabetes beta cell function (Morris et al., 2012[85]) | Alzheimer's Disease (Lambert et al., 2013[49]) | 9 | NR | OR=1.00 95% CI:0.94;1.07 |
| | | Type 2 diabetes insulin sensitivity (Morris et al., 2012[85]) | Alzheimer's Disease (Lambert et al., 2013[49]) | 2 | NR | OR=1.17 95% CI:1.02;1.34* |
| | | Type 2 diabetes other biological factors (Morris et al., 2012[85]) | Alzheimer's Disease (Lambert et al., 2013[49]) | 10 | NR | OR=0.90 95% CI:0.79;1.04 |
| | Zhang et al., (2020)[42] | Type 2 diabetes (Scott et al., 2017[81]) | Alzheimer's Disease (Lambert et al., 2013[49]) | 101 | NR | OR=1.01 95% CI:0.96;0.63 |
| | | Fasting insulin (Scott et al., 2017[81]) | Alzheimer's Disease (Lambert et al., 2013[49]) | 13 | NR | OR=1.50 95% CI:0.86;2.60 |
| | | Fasting glucose (Scott et al., 2017[81]) | Alzheimer's Disease (Lambert et al., 2013[49]) | 32 | NR | OR=1.30 95% CI:1.01;1.66* |
| | Zhou et al., (2019)[44] | Fasting insulin (Scott et al., 2017[81]) | Alzheimer's Disease (Jansen et al., 2019[57]) | 12 | 1016 | OR=1.13 95% CI:1.04;1.23** |
| | | Insulin resistance (Knowles et al., 2015[90]) | Alzheimer's Disease (Jansen et al., 2019[57]) | 5 | 72 | OR=1.02 95% CI:1.00;1.03** |
| | Xue et al., (2023)[40] | Type 2 diabetes (Mahajan et al., 2018[83]) | Alzheimer's Disease (Kunkle et al., 2019[51]) | 53 | >10 | OR=1.24 95% CI:1.10;1.39** |
| | Andrews et al., (2021)[4] | Social isolation (Day et al., 2018[91]) | Alzheimer's Disease (Kunkle et al., 2019[51]) | 14 | 39.0 | $\beta$=-0.42 SE=0.31 |

| Modifiable Risk Factor | Study | Exposure (GWAS dataset/consortium) | Outcome (GWAS dataset/consortium) | N SNPs | *F*-statistic | IVW Estimate |
|---|---|---|---|---|---|---|
| **Social contact related exposures** | Desai et al., (2023)[8] | Social isolation (Day et al., 2018[91]) | Alzheimer's Disease and Alzheimer's Disease by Proxy (Bellenguez et al., 2023[52]) | 15 | 36.1 | OR=1.23 95% CI:0.76;1.98 |
| | | Social isolation (Day et al., 2018[91]) | Dementia with Lewy Bodies (Chia et al., 2021[53]) | 14 | 35 | OR=0.22 95% CI:0.03;1.47 |
| | | Social isolation (Day et al., 2018[91]) | Frontotemporal Dementia (Ferrari et al., 2014[54]) | 14 | 36.4 | OR=3.80 95% CI:0.68;21.30 |
| | Shen et al., (2021)[32] | Regular pub (Day et al., 2018[91]) | Alzheimer's Disease (Jansen et al., 2019[57]) | 11 | 44.4 | OR=1.19 95% CI:0.98;1.44 |
| | | Regular pub (Day et al., 2018[91]) | Alzheimer's Disease by proxy (Marioni et al., 2018[55]) | 11 | 44.4 | OR=0.97 95% CI:0.51;1.85 |
| | | Regular gym (Day et al., 2018[91]) | Alzheimer's Disease (Jansen et al., 2019[57]) | 6 | 82.5 | OR=0.67 95% CI:0.46;0.97 |
| | | Regular gym (Day et al., 2018[91]) | Alzheimer's Disease by proxy (Marioni et al., 2018[55]) | 5 | NR | OR=0.16 95% CI:0.04;0.62* |
| | | Regular religious group (Day et al., 2018[91]) | Alzheimer's Disease (Jansen et al., 2019[57]) | 18 | 40.3 | OR=0.85 95% CI:0.68;1.07 |
| | | Regular religious group (Day et al., 2018[91]) | Alzheimer's Disease by proxy (Marioni et al., 2018[55]) | 14 | NR | OR=0.62 95% CI:0.21;1.81 |
| | | Loneliness (Day et al., 2018[91]) | Alzheimer's Disease (Jansen et al., 2019[57]) | 13 | 90.2 | OR=0.90 95% CI:0.63;1.29 |
| | | Loneliness (Day et al., 2018[91]) | Alzheimer's Disease by proxy (Marioni et al., 2018[55]) | 11 | NR | OR=0.89 95% CI:0.23;3.42 |
| | | Loneliness (Day et al., 2018[91]) | Alzheimer's Disease (Jansen et al., 2019[57]) | 15 | 99.1 | OR=0.99 95% CI:0.87;0.91 |
| | | Loneliness (Day et al., 2018[91]) | Alzheimer's Disease by proxy (Marioni et al., 2018[55]) | 14 | 99.1 | OR=0.95 95% CI:0.66;1.37 |
| | Thorp et al., (2022)[35] | Loneliness (Day et al., 2018[91]) | Alzheimer's Disease (Lambert et al., 2013[49]) | 10 | 35.9 | OR=1.12 95% CI:0.13;9.74 |
| | Andrews et al., (2021)[4] | Moderate-to-vigorous physical activity (Klimentidis et al., 2018[92]) | Alzheimer's Disease (Kunkle et al., 2019[51]) | 20 | 33.3 | $\beta$=0.17 SE=0.25 |

| Modifiable Risk Factor | Study | Exposure (GWAS dataset/consortium) | Outcome (GWAS dataset/consortium) | N SNPs | *F*-statistic | IVW Estimate |
|---|---|---|---|---|---|---|
| **Physical activity related exposures** | Baumeister et al., (2020)[5] | Average physical activity (Klimentidis et al., 2018[92]) | Alzheimer's Disease (Kunkle et al., 2019[51]) | 8 | >30 | OR=1.03 95% CI:0.97;1.10 |
| | | Vigorous physical activity (Klimentidis et al., 2018[92]) | Alzheimer's Disease (Kunkle et al., 2019[51]) | 8 | >25 | OR=0.91 95% CI:0.46;1.81 |
| | | Average physical activity (Klimentidis et al., 2018[92]) | Alzheimer's Disease (Jansen et al., 2019[57]) | 8 | >30 | OR=0.99 95% CI:0.99;1.01 |
| | | Vigorous physical activity (Klimentidis et al., 2018[92]) | Alzheimer's Disease (Jansen et al., 2019[57]) | 8 | >25 | OR=1.04 95% CI:0.95;1.15 |
| | Chen et al., (2022)[6] | Time spent using a computer (MRC-IEU consortium) | Alzheimer's Disease (Kunkle et al., 2019[51]) | 76 | 39 | OR=0.71 95% CI:0.52;0.99* |
| | Desai et al., (2023)[8] | Vigorous physical activity (Klimentidis et al., 2018[92]) | Alzheimer's Disease and Alzheimer's Disease by Proxy (Bellenguez et al., 2023[52]) | 7 | 38 | OR=0.48 95% CI:0.10;2.30 |
| | | Vigorous physical activity (Klimentidis et al., 2018[92]) | Dementia with Lewy Bodies (Chia et al., 2021[53]) | 7 | 38 | OR=0.007 95% CI:>.0001;10.45 |
| | | Vigorous physical activity (Klimentidis et al., 2018[92]) | Frontotemporal Dementia (Ferrari et al., 2014[54]) | 7 | 38.2 | OR=4.57 95% CI:0.12;180.71 |
| | He et al., (2022)[17] | Sedentary behaviours (TV watching) (van de Vegte et al., 2020[93]) | Alzheimer's Disease (Kunkle et al., 2019[51]) | 85 | >30 | OR=1.50 95% CI:0.91;1.46 |
| | | Sedentary behaviours (computer use) (van de Vegte et al., 2020[93]) | Alzheimer's Disease (Kunkle et al., 2019[51]) | 22 | >30 | OR=0.52 95% CI:0.32;0.84* |
| | | Sedentary behaviours (driving) (van de Vegte et al., 2020[93]) | Alzheimer's Disease (Kunkle et al., 2019[51]) | 4 | >30 | OR=0.64 95% CI:0.22;1.92 |
| | Korologou-Linden et al., (2022)[14] | Self-reported moderate physical activity (UK Biobank) | Alzheimer's Disease (Lambert et al., 2013[49]) | 14 | >30 | OR=1.40 95% CI:1.06;1.85* |
| | Liao et al., (2022)[17] | Fraction of accelerations >425 milli-gravities (Klimentidis et al., 2018[92]) | Alzheimer's Disease (Kunkle et al., 2019[51]) | 2 | NR | OR=0.39 95% CI:0.08;1.98 |
| | | Overall acceleration average (Klimentidis et al., 2018[92]) | Alzheimer's Disease (Kunkle et al., 2019[51]) | 8 | NR | OR=0.99 95% CI:0.77;1.27 |

| Modifiable Risk Factor | Study | Exposure (GWAS dataset/consortium) | Outcome (GWAS dataset/consortium) | N SNPs | *F*-statistic | IVW Estimate |
|---|---|---|---|---|---|---|
| | | Moderate-to-vigorous physical activity (Klimentidis et al., 2018[92]) | Alzheimer's Disease (Kunkle et al., 2019[51]) | 9 | NR | OR=0.75 95% CI:0.20;2.75 |
| | | Vigorous physical activity (Klimentidis et al., 2018[92]) | Alzheimer's Disease (Kunkle et al., 2019[51]) | 5 | NR | OR=0.83 95% CI:0.08;8.53 |
| | Malik et al., (2021)[21] | Overall physical activity (UK Biobank) | Incident Dementia (UK Biobank) | 3 | NR | OR=1.59 95% CI:0.14;18.30 |
| | Thorp et al., (2022)[35] | Physical activity (Klimentidis et al., 2018[92]) | Alzheimer's Disease (Lambert et al., 2013[49]) | 18 | 33.4 | OR=0.93 95% CI:0.52;1.66 |
| | Wu et al., (2021)[39] | Physical activity (Doherty et al., 2018[94]) | Alzheimer's Disease (Kunkle et al., 2019[51]) | 5 | NR | OR=1.03 95% CI:0.48;2.21 |
| | | Physical activity (Klimentidis et al., 2018[92]) | Alzheimer's Disease (Kunkle et al., 2019[51]) | 8 | NR | OR=1.03 95% CI:0.96;1.10 |
| | Yang et al., (2021)[41] | Sedentary behaviours (TV watching) (van de Vegte et al., 2020[93]) | Alzheimer's Disease (Kunkle et al., 2019[51]) | NR | 5747 | OR=1.15 95% CI:0.97;1.36 |
| | | Sedentary behaviours (computer use) (van de Vegte et al., 2020[93]) | Alzheimer's Disease (Kunkle et al., 2019[51]) | NR | 1685 | OR=0.67 95% CI:0.48;0.92* |
| | | Sedentary behaviours (driving) (van de Vegte et al., 2020[93]) | Alzheimer's Disease (Kunkle et al., 2019[51]) | NR | 193 | OR=1.26 95% CI:0.50;3.19 |
| | Zhang et al., (2020)[42] | Overall activity (Doherty et al., 2018[94]) | Alzheimer's Disease (Kunkle et al., 2019[51]) | 3 | 18.2 | OR=0.62 95% CI:0.17;2.32 |
| | | Sedentary behaviour (Doherty et al., 2018[94]) | Alzheimer's Disease (Kunkle et al., 2019[51]) | 3 | 18.2 | OR=1.33 95% CI:0.68;2.60 |
| | | Walking (Doherty et al., 2018[94]) | Alzheimer's Disease (Kunkle et al., 2019[51]) | 2 | 27.4 | OR=0.30 95% CI:0.13;0.68* |
| | | Moderate-intensity activity (Doherty et al., 2018[94]) | Alzheimer's Disease (Kunkle et al., 2019[51]) | 25 | 2.19 | OR=0.80 95% CI:0.59;1.07 |
| | Andrews et al., (2021)[4] | Cigarettes per day (Liu et al., 2019[60]) | Alzheimer's Disease (Kunkle et al., 2019[51]) | 43 | 89.7 | $\beta$=-0.03 SE=0.15 |
| | | Smoking initiation (Liu et al., 2019[60]) | Alzheimer's Disease (Kunkle et al., 2019[51]) | 342 | 51.2 | $\beta$=-0.004 SE=0.14 |

| Modifiable Risk Factor | Study | Exposure (GWAS dataset/consortium) | Outcome (GWAS dataset/consortium) | N SNPs | *F*-statistic | IVW Estimate |
|---|---|---|---|---|---|---|
| **Smoking related exposures** | Desai et al., (2023)[8] | Lifetime smoking (Wootton et al., 2019[95]) | Alzheimer's Disease and Alzheimer's Disease by Proxy (Bellenguez et al., 2023[52]) | 126 | 42.7 | OR=0.80 95% CI:0.69;0.92* |
| | | Lifetime smoking (Wootton et al., 2019[95]) | Dementia with Lewy Bodies (Chia et al., 2021[53]) | 119 | 42.8 | OR=0.83 95% CI:0.49;1.42 |
| | | Lifetime smoking (Wootton et al., 2019[95]) | Frontotemporal Dementia (Ferrari et al., 2014[54]) | 123 | 88.4 | OR=0.71 95% CI:0.13;3.98 |
| | Huang et al., (2023)[13] | Smoking initiation (Liu et al., 2019[60]) | Alzheimer's Disease (Kunkle et al., 2019[51]) | 73 | NR | OR=0.95 95% CI:0.81;1.13 |
| | | Smoking initiation (Liu et al., 2019[60]) | Alzheimer's Disease by Proxy - Maternal history (Marioni et al., 2018[55]) | 73 | NR | OR=0.96 95% CI:0.81;1.14 |
| | | Smoking initiation (Liu et al., 2019[60]) | Alzheimer's Disease by Proxy - Paternal history (Marioni et al., 2018[55]) | 73 | NR | OR=0.91 95% CI:0.68;1.23 |
| | | Cigarettes per day (Liu et al., 2019[60]) | Alzheimer's Disease (Kunkle et al., 2019[51]) | 20 | NR | OR=0.96 95% CI:0.83;1.12 |
| | | Cigarettes per day (Liu et al., 2019[60]) | Alzheimer's Disease by Proxy - Maternal history (Marioni et al., 2018[55]) | 20 | NR | OR=1.18 95% CI:1.01;1.37* |
| | | Cigarettes per day (Liu et al., 2019[60]) | Alzheimer's Disease by Proxy - Paternal history (Marioni et al., 2018[55]) | 20 | NR | OR=1.09 95% CI:0.89;1.25 |
| | Malik et al., (2021)[21] | Smoking index (UK Biobank) | Incident Dementia (UK Biobank) | 126 | NR | OR=1.22 95% CI:0.77;1.94 |
| | Larsson et al., (2017)[15] | Smoking quantity (Thorgeirsson et al., 2010[96]) | Alzheimer's Disease (Lambert et al., 2013[49]) | 4 | 68 | OR=0.69 95% CI:0.49;0.99* |
| | | Smoking initiation (Furberg et al., 2010[97]) | Alzheimer's Disease (Lambert et al., 2013[49]) | 1 | 16 | OR=0.71 95% CI:0.37;1.33 |
| | | Smoking cessation (Furberg et al., 2010[97]) | Alzheimer's Disease (Lambert et al., 2013[49]) | 1 | 103 | OR=1.16 95% CI:0.75;1.78 |
| | Luo et al., (2023)[20] | Smoking regularly (Liu et al., 2019[60]) | Alzheimer's Disease (Bellenguez et al., 2022[52]) | 313 | >10 | OR=0.93 95% CI:0.87;1.00 |

| Modifiable Risk Factor | Study | Exposure (GWAS dataset/consortium) | Outcome (GWAS dataset/consortium) | N SNPs | *F*-statistic | IVW Estimate |
|---|---|---|---|---|---|---|
| | | Smoking regularly (Liu et al., 2019[60]) | Alzheimer's Disease by proxy (Bellenguez et al., 2022[52]) | 312 | >10 | OR=0.88 95% CI:0.82;0.82* |
| | Zhang et al., (2020)[42] | Current smoking (UK Biobank) | Alzheimer's Disease (Lambert et al., 2013[49]) | 27 | NR | OR=0.74 95% CI:0.32;1.69 |
| | Zhu et al., (2023)[46] | Cigarettes per day (Matoba et al., 2019[98]) | Alzheimer's Disease Chinese Cohort (Zhu et al., 2023[46]) | 5 | 53 | OR=0.51 95% CI:0.15;1.74 |
| | | Cigarettes per day (Matoba et al., 2019[98]) | Alzheimer's Disease Japanese Cohort (Shigemizu et al., 2021[99]) | 5 | 53 | OR=1.70 95% CI:0.79;1.73 |
| | Østergaard et al., (2015)[28] | Smoking quantity (Furberg et al., 2010[97]) | Alzheimer's Disease (Lambert et al., 2013[49]) | 3 | NR | OR=0.67 95% CI:0.51;0.89* |
| | | Smoking initiation (Furberg et al., 2010[97]) | Alzheimer's Disease (Lambert et al., 2013[49]) | 1 | NR | OR=0.70 95% CI:0.37;1.33 |
| | Nordesthaard et al., (2022)[27] | Smoking cumulative (CGPS & CCHS) | All-cause Dementia (CGPS & CCHS) | 1 | 74 | OR=1.04 95% CI:0.96;1.11 |
| | | Smoking cumulative (CGPS & CCHS) | Alzheimer's Disease (CGPS & CCHS) | 1 | 74 | OR=1.06 95% CI:0.97;1.16 |
| | | Smoking cumulative (CGPS & CCHS) | Non-Alzheimer's dementia (CGPS & CCHS) | 1 | 74 | OR=0.98 95% CI:0.88;1.10 |
| | | Smoking cumulative (CGPS & CCHS) | Alzheimer's Disease (Lambert et al., 2013[49]) | 1 | 74 | OR=0.22 95% CI:0.18;1.28 |
| | Thorp et al., (2022)[35] | Smoking initiation (Liu et al., 2019[60]) | Alzheimer's Disease (Lambert et al., 2013[49]) | 77 | 41.8 | OR=0.92 95% CI:0.77;1.09 |
| | | Cigarettes per day (Liu et al., 2019[60]) | Alzheimer's Disease (Lambert et al., 2013[49]) | 20 | 104 | OR=0.98 95% CI:0.83;1.15 |

GWAS: Genome Wide Association Study; IVW: Inverse variance weighted; SNP: Single Nucleotide Polymorphism; CI: Confidence Interval; OR: Odds Ratio; $\beta$: beta; SE: Standard Error; NR: Not Reported; *$p$<.05; **$p$<.001;***$p$<.0001; AUDIT: Alcohol Use Disorders Identification Test; HR: Hazards Ratio; CGPS: Copenhagen General Population Study; CCHS: Copenhagen City Heart Study
